# Differences in Dietary Intake Exist Among U.S. Adults by Diabetic Status Using NHANES 2009-2016

**DOI:** 10.1101/2022.05.18.22275288

**Authors:** Luotao Lin, Fengqing Zhu, Edward J. Delp, Heather A. Eicher-Miller

**Author notes:** **Correspondence:** Heather Eicher-Miller.

## Abstract

**Objectives:** Insulin takers’ diets have rarely been described although insulin dosing is highly dependent on dietary intake. The objective of this paper was to determine the most frequently consumed food or beverage items, food subcategories, and food categories, and those that contributed most to total energy intake among U.S. insulin takers, those with type 2 diabetes (T2D) not taking insulin and those without diabetes; the groups were also compared at the broad food category level.

**Methods:** The Laboratory tests and questionnaires of the NHANES 2009–2016 classified 774 insulin takers, 2,758 T2D not taking insulin, and 17,796 participants without diabetes. USDA assigned 8-digit food codes identifying each food item and its membership to a category and subcategory of foods were used to categorize foods based on the WWEIA categories. Raw and weighted frequency and energy contributions of each food item, food subcategory, and food category were calculated and ranked. Comparisons among groups by broad food category used the Rao–Scott modified chi-square test.

**Results:** Diet soft drinks ranked as the 4^th^ and 7^th^ most consumed food subcategory in insulin takers and T2D not taking insulin, respectively. Soft drinks ranked as the 8^th^ and 6^th^ most consumed food subcategory in T2D not taking insulin and those without diabetes, and contributed 5^th^ and 2^nd^ most to energy, respectively. Protein foods represented 4 of the top 10 highest energy contributing food subcategories among insulin takers, 3 of the top 10 food subcategories among those with T2D not taking insulin, and only 1 subcategory among those without diabetes. Insulin takers had higher consumption frequency of grains, and lower consumption frequency of sweets and alcohol, and a larger share of energy comprising protein, vegetables, and grains, and a smaller share of energy comprising beverages and alcohol compared to participants without diabetes.

**Conclusions:** Differences in dietary intake exist among U.S. adults by diabetic status. Insulin takers are likely to consume more protein foods and less regular soft drinks compared to other 2 groups. Lists of the most frequently reported foods and foods contributing most to energy may be helpful for nutrition education, prescribing diets, and digital-based dietary assessment for insulin takers.

## 1 Introduction

Diabetes is a chronic disease highly influenced by diet where individuals either do not produce enough insulin to regulate blood glucose level or they are not able to use the insulin they produce to effectively move blood glucose into body cells (1). Over time, diabetes can cause additional serious health problems including heart disease, vision loss, and kidney disease. About 34.2 million U.S. individuals (10.5% of the U.S. population) diabetes including type 1 (T1D), type 2 (T2D), and gestational diabetes in 2020 (2).

Both those with T1D and T2D may use injectable or oral insulin to control their blood glucose levels. Proper insulin dosing to control blood glucose levels and reduce risk of severe consequences is dependent on dietary intake, particularly the frequency and energy from macronutrients: carbohydrate, protein, and fat (3,4). Thus, quantification of the foods comprising dietary intake may be used to inform insulin dosing algorithms to enhance blood glucose level control (5,6) and knowledge of the foods comprising overall dietary intake may also aid the creation of meal plans, inform substitutions, and other interventions. Furthermore, it is likely that diabetes status may differentiate food and beverage choice and intake among those using insulin, T2D not using insulin, and those without diabetes as one strategy for glucose control is through the avoidance of certain foods and monitored intake of carbohydrates, for example sugar-sweetened soft drinks (7). However, very little information on the food intake of those taking insulin or those with T2D not taking insulin is available (8–10).

Therefore, the objectives of this study were to (1) determine the most frequently consumed food items, food subcategories, and food categories and the food items, food subcategories, and food categories that contributed most to energy intake, and (2) compare the differences of the most frequently consumed broad food categories and the broad food categories that contributed most to energy intake among insulin takers, T2D participants not taking insulin, and participants without diabetes among U.S. adults aged 18 years old or older using data from the National Health And Nutrition Examination Survey (NHANES) 2009–2016.

## 2 Materials and methods

### 2.1 Participants and Dataset

NHANES is a cross-sectional survey that is conducted by the National Center for Health Statistics (NCHS) of the U.S. Centers for Disease Control and Prevention to assess the health and nutritional status of the non-institutionalized civilian population in the U.S. (11). Participants’ sociodemographic information was obtained via questionnaires during the in-person household interview, which included age, sex, race/ethnicity, and poverty to income ratio (PIR). The NCHS Research Ethics Review Board approved the survey and all participants consented to participate (12).

### 2.2 Analytic Sample

NHANES 2009-2016 was used to include the most recent available dietary data and sufficient sample size. Adults in the United States aged 18 or older who had a reliable first 24-hour dietary recall, reliable laboratory test results, and valid answers to diabetic-related questions made up the analytic sample.

NHANES diabetic-related questions and laboratory test results were used to divide the sample into three groups: insulin takers, participants with diabetes not taking insulin, and participants without diabetes. The number of insulin takers was determined by the question “Are you/ is the sample person now taking insulin?” Those participants with diabetes who were not taking insulin were identified by subtracting those using insulin from the total number of participants with diabetes. Diabetes was classified by self-reporting being told they had diabetes by a doctor or taking medications that lowered glucose, or by fasting plasma glucose concentration ≥ 126 mg/dL or hemoglobin A1c ≥ 6.5% (13). Since T1D was classified when a participant reported being diagnosed with diabetes before the age of 30 years old and reporting continuous insulin use since diagnosis (14), insulin takers encompass all those with T1D, so participants with diabetes not taking insulin is equal to participants with T2D not taking insulin. Those participants without diabetes were identified by subtracting the number of participants with diabetes from the sample. As a result, the study sample included 774 insulin takers, 2,758 T2D who did not use insulin, and 17,796 participants who did not have diabetes.

### 2.3 Sociodemographic characteristics

Sociodemographic characteristics included survey year (2009-2010, 2011-2012, 2013-2014, and 2015-2016) and self-reported sex (male or female), race/ethnicity (grouped as Hispanic, non-Hispanic white, non-Hispanic black, and other including multi-race), age (grouped as 18-34, 35-49, and 50-80 years), and PIR which was the reported household income divided by the federal poverty guideline for household income, and grouped as 0-0.99 (below poverty threshold), 1-1.99, 2-2.99, 3-3.99, and 4- 5 (15).

### 2.4 Dietary Assessment

The USDA Automated Multiple-Pass Method (16) was used to collect a dietary recall recorded during the physical health examination where participants reported all of the foods consumed during the previous 24-hour period, including information on the time of intake, amount and type of each food, and detailed food descriptions (17). Each reported food item was then linked to the USDA’s Food and Nutrient Database for Dietary Studies (FNDDS) (18–21) to assign an 8-digit food codes. The USDA food codes were also used to sort the reported foods into the What We Eat in America food categories and subcategories (22).

### 2.5 Statistical Analysis

The Rao-Scott modified chi-square test was used to compare sociodemographic variables created from the survey data among the three groups.

The weighted frequencies of reported foods were computed by determining the weighted sum of the number of food items, food subcategories, and food categories:

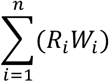

where n= the sample size, i= each participant, *R*_*i*_ = # of reports of individual food code for the i^th^ participant, *W*_*i*_= dietary day 1 sample weight for the i^th^ participant (23,24). The number of reports of foods for each participant was multiplied by the dietary day 1 sample weight, and then summed to yield the total weighted frequency of the particular food or beverage for each respective group, which represented how often that food or beverage was consumed by the respective group in the U.S. in one day. The weighted frequency of the food or beverage was also divided by the total weighted frequency of all foods or beverages reported in a single day to get the weighted percent of the frequency of that food or beverage being consumed out of all reported foods and beverages consumed.

The weighted energy contribution of reported foods items, food subcategories, and food categories were computed by determining the weighted sum as:

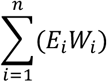

where n= the sample size, i= each participant *E*_*i*_ =total energy for an individual food code for the i^th^ participant, *W*_*i*_ = dietary day 1 sample weight for the i^th^ participant (23,24). The energy contributed by a particular food or beverage was first summed for each participant and then multiplied by the dietary day 1 sample weight of the participant, then summed for all participants in the sample for each group, which represented how much energy that food or beverage contributed to the total energy for the respective group in the U.S. in one day. The weighted energy of the particular food or beverage was also divided by the total weighted energy from all reported foods and beverages to get the weighted percent of energy from that food or beverage out of the energy estimated from all reported foods and beverages.

The Rao-Scott modified chi-square goodness-of-fit tests were used to compare the frequency and energy contribution comparisons of food terms among the three groups. Significantly differences were indicated as when p < 0.05/42 or p < 0.0012 using a Bonferroni type adjustment for multiple comparisons for food intake among 14 broad food categories × 3 groups. Survey weights, or the reciprocal of sample inclusion probability, were used in all computations to allow for inference to the non-institutionalized U.S. population. Further adjustments were performed to account for the clustering and stratification inherent to the survey design. All analyses were completed in SAS 9.4 (SAS Institute Inc).

## 3 Results

### 3.1 Sociodemographic characteristics

The sociodemographic characteristics of participants taking insulin, with T2D not taking insulin, and participants without diabetes are shown in **Table 1**. Sex, age, race/ethnicity, and PIR were significantly different among the three groups.

**Table 1.** Sociodemographic characteristics of U.S. adults 18 y or older taking insulin, with Type 2 diabetes but not taking insulin, and without diabetes as drawn from the NHANES, 2009-2016^1^.

|  | Insulin takers<br>(n=774) | T2D w/o insulin<br>(n=2,758) | Participants w/o<br>diabetes<br>(n=17,796) | P-value<sup>2</sup> |
| --- | --- | --- | --- | --- |
| Sex |  |  |  | 0.0003* |
| Male | 403 (52.1%) | 1416 (51.3%) | 8578 (48.2%) |  |
| Female | 371 (47.9%) | 1342 (48.7%) | 9218 (51.8%) |  |
| Age in Years |  |  |  | <0.0001* |
| 18-34 | 44 (5.6%) | 121 (4.4%) | 6247 (35.1%) |  |
| 35-49 | 91 (11.8%) | 470 (17.0%) | 4633 (26.0%) |  |
| 50-80 | 639 (82.6%) | 2167 (78.6%) | 6916 (38.9%) |  |
| Survey year |  |  |  | 0.4 |
| 2009-2010 | 193 (24.9%) | 766 (27.8%) | 5005 (28.1%) |  |
| 2011-2012 | 181 (23.4%) | 647 (23.5%) | 4168 (23.4%) |  |
| 2013-2014 | 181 (23.4%) | 660 (23.9%) | 4387 (24.7%) |  |
| 2015-2016 | 219 (28.3%) | 685 (24.8%) | 4236 (23.8%) |  |
| Race/Ethnicity |  |  |  | <0.0001* |
| Hispanic | 205 (27.7%) | 859 (31.1%) | 4448 (25.0%) |  |
| White | 276 (35.7%) | 905 (32.8%) | 7517 (42.2%) |  |
| Black | 229 (29.6%) | 692 (25.1%) | 3640 (20.5%) |  |
| Other | 54 (7.0%) | 302 (11.0%) | 2191 (12.3%) |  |
| Poverty Income<br>Ratio |  |  |  | <0.0001* |
| 0.00-0.99 | 201 (28.1%) | 623 (25.0%) | 3776 (23.2%) |  |
| 1.00-1.99 | 220 (30.7%) | 748 (30.0%) | 4219 (25.9%) |  |
| 2.00-2.99 | 106 (14.8%) | 384 (15.4%) | 2353 (14.4%) |  |
| 3.00-3.99 | 66 (9.2%) | 277 (11.1%) | 1793 (11.0%) |  |
| 4.00-5.00 | 123 (17.2%) | 463 (18.5%) | 4156 (25.5%) |  |

<sup>1</sup> Total numbers do not always add to sample size due to missing values. Percentages do not always add to 100 due to rounding.
<sup>2</sup> Rao–Scott modified chi-square p-value is shown. Analyses were adjusted for survey design including stratification and clustering. Sample weights were appropriately constructed and applied to this analysis as directed by NCHS. Weights were rescaled so that the sum to the weights matched the survey population at the midpoint of the 8 years, 2009–2016. Statistical significance for differences among three groups when $p < 0.05$ is indicated by “\*”.

Most insulin takers and those with T2D who were not using insulin were ages 50 to 80 years. Participants without diabetes were more evenly distributed in three age groups (18-34, 35-49, 50-80). About half of the group using insulin and with T2D but not taking insulin had PIR <2, furthermore, 28.1% of insulin takers had PIR below the poverty threshold (PIR<1). The group without diabetes had higher incomes with more than 25% of participants having a PIR greater than 4.

### 3.2 The most frequently consumed food items, food subcategories, and food categories

The top 10 most frequently reported food items, food subcategories, and food categories for insulin takers, those with T2D not using insulin, and participants without diabetes are shown in **Tables 2-4**. Foods are presented in descending weighted frequency order by their FNDDS short food descriptions. In frequency lists, 7 food items (Table 2), 6 food subcategories (Table 3), and 8 food categories (Table 4) are common across the three groups but their rankings were varied. Notably, diet soft drink was the 4^th^ most frequently consumed food subcategory among insulin takers, the 7^th^ among those with T2D not taking insulin, and did not appear in the top 10 food subcategory frequency list among those without diabetes (Table3). Regular soft drink did not appear in the top 10 food subcategory frequency list for insulin takers, but was the 8^th^ most frequently consumed food subcategory among those with T2D not taking insulin, and the 6^th^ most frequently consumed food subcategory among participants without diabetes (Table3).

**Table 2.**
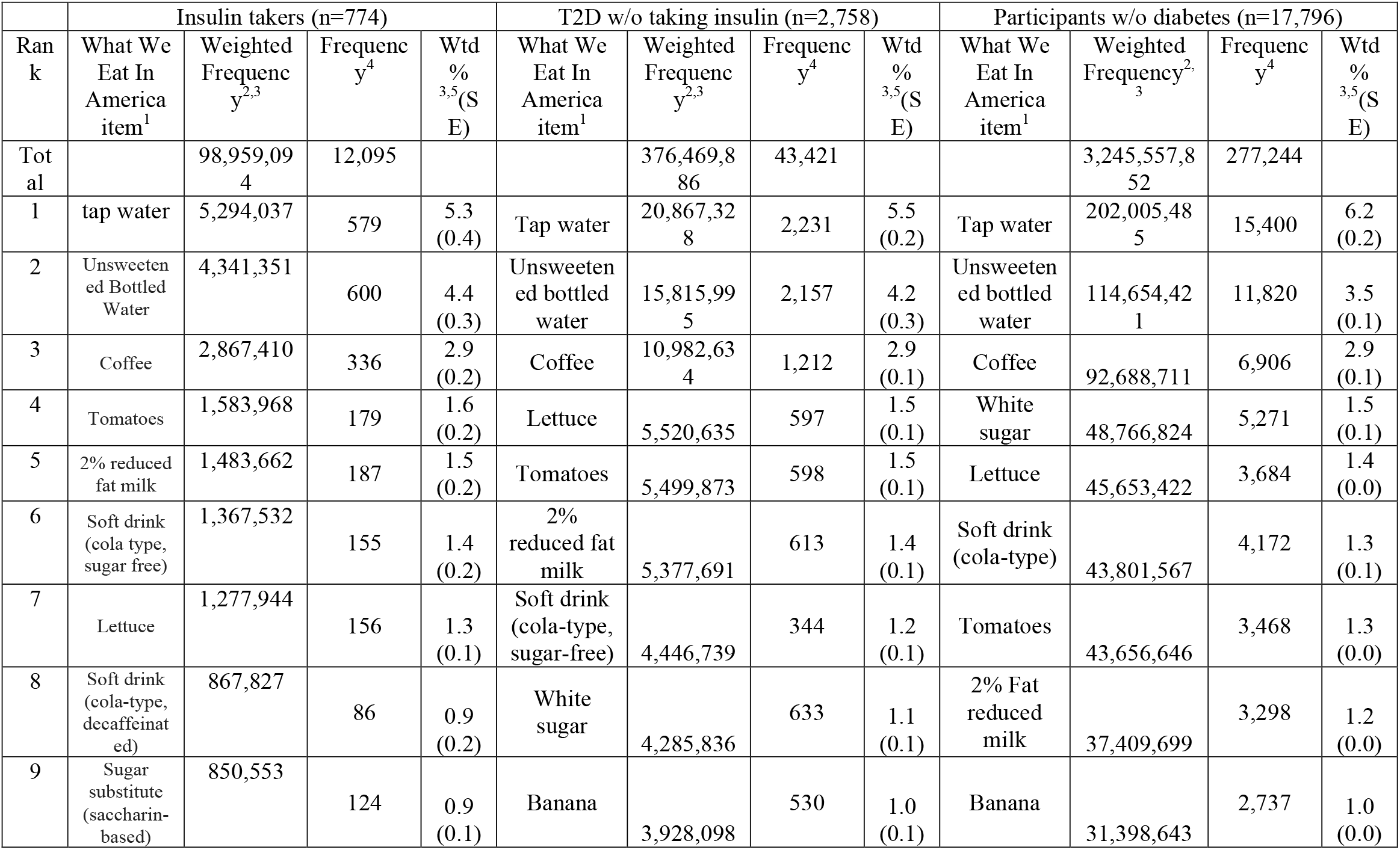
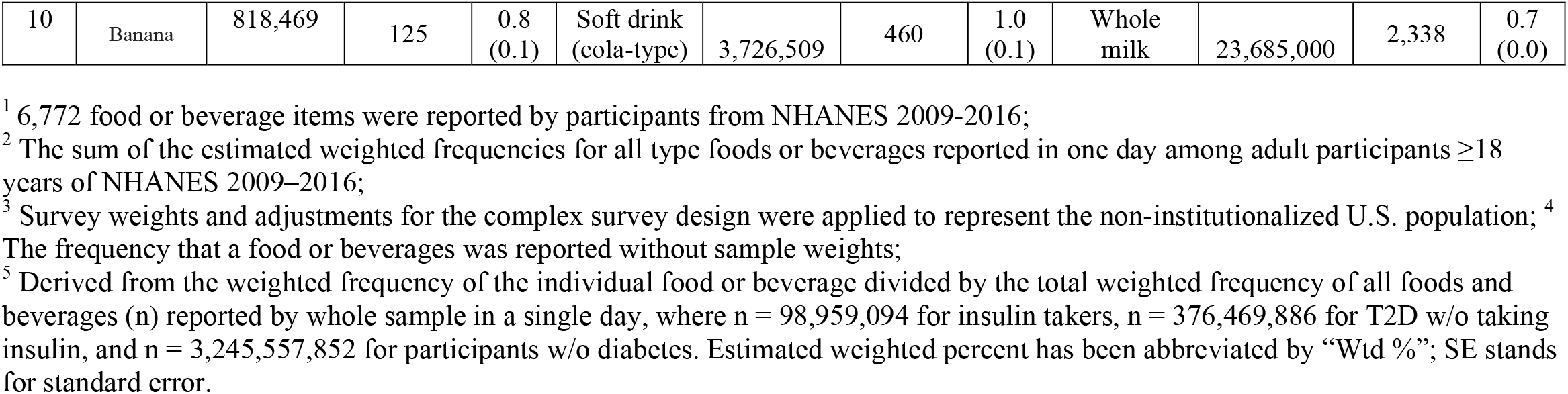
Top 10 most frequently consumed foods items or beverages^1^, unweighted and weighted frequency of reported foods items or beverages, percent of reported foods or beverages, and standard error of percent of reported foods or beverages among all reported foods or beverages for U.S. adults 18 y or older taking insulin, with Type 2 diabetes but not taking insulin, and without diabetes drawn from 2009–2016 NHANES data.

|  | Insulin takers (n=774) |  |  |  | T2D w/o taking insulin (n=2,758) |  |  |  | Participants w/o diabetes (n=17,796) |  |  |  |
| --- | --- | --- | --- | --- | --- | --- | --- | --- | --- | --- | --- | --- |
| Ran<br>k | What We<br>Eat In<br>America<br>item<sup>1</sup> | Weighted<br>Freque<br>nc<br>y<sup>2,3</sup> | Freque<br>nc<br>y<sup>4</sup> | Wtd<br>%<br>3,5(S<br>E) | What We<br>Eat In<br>America<br>item<sup>1</sup> | Weighted<br>Freque<br>nc<br>y<sup>2,3</sup> | Freque<br>nc<br>y<sup>4</sup> | Wtd<br>%<br>3,5(S<br>E) | What We<br>Eat In<br>America<br>item<sup>1</sup> | Weighted<br>Frequency<sup>2,3</sup> | Freque<br>nc<br>y<sup>4</sup> | Wtd<br>%<br>3,5(S<br>E) |
| Tot<br>al |  | 98,959,094 | 12,095 |  |  | 376,469,886 | 43,421 |  |  | 3,245,557,852 | 277,244 |  |
| 1 | tap water | 5,294,037 | 579 | 5.3<br>(0.4) | Tap water | 20,867,328 | 2,231 | 5.5<br>(0.2) | Tap water | 202,005,485 | 15,400 | 6.2<br>(0.2) |
| 2 | Unsweeten<br>ed Bottled<br>Water | 4,341,351 | 600 | 4.4<br>(0.3) | Unsweeten<br>ed bottled<br>water | 15,815,995 | 2,157 | 4.2<br>(0.3) | Unsweeten<br>ed bottled<br>water | 114,654,421 | 11,820 | 3.5<br>(0.1) |
| 3 | Coffee | 2,867,410 | 336 | 2.9<br>(0.2) | Coffee | 10,982,634 | 1,212 | 2.9<br>(0.1) | Coffee | 92,688,711 | 6,906 | 2.9<br>(0.1) |
| 4 | Tomatoes | 1,583,968 | 179 | 1.6<br>(0.2) | Lettuce | 5,520,635 | 597 | 1.5<br>(0.1) | White<br>sugar | 48,766,824 | 5,271 | 1.5<br>(0.1) |
| 5 | 2% reduced<br>fat milk | 1,483,662 | 187 | 1.5<br>(0.2) | Tomatoes | 5,499,873 | 598 | 1.5<br>(0.1) | Lettuce | 45,653,422 | 3,684 | 1.4<br>(0.0) |
| 6 | Soft drink<br>(cola type,<br>sugar free) | 1,367,532 | 155 | 1.4<br>(0.2) | 2%<br>reduced fat<br>milk | 5,377,691 | 613 | 1.4<br>(0.1) | Soft drink<br>(cola-type) | 43,801,567 | 4,172 | 1.3<br>(0.1) |
| 7 | Lettuce | 1,277,944 | 156 | 1.3<br>(0.1) | Soft drink<br>(cola-type,<br>sugar-free) | 4,446,739 | 344 | 1.2<br>(0.1) | Tomatoes | 43,656,646 | 3,468 | 1.3<br>(0.0) |
| 8 | Soft drink<br>(cola-type,<br>decaffeinat<br>ed) | 867,827 | 86 | 0.9<br>(0.2) | White<br>sugar | 4,285,836 | 633 | 1.1<br>(0.1) | 2% Fat<br>reduced<br>milk | 37,409,699 | 3,298 | 1.2<br>(0.0) |
| 9 | Sugar<br>substitute<br>(saccharin-<br>based) | 850,553 | 124 | 0.9<br>(0.1) | Banana | 3,928,098 | 530 | 1.0<br>(0.1) | Banana | 31,398,643 | 2,737 | 1.0<br>(0.0) |
| 10 | Banana | 818,469 | 125 | 0.8<br>(0.1) | Soft drink<br>(cola-type) | 3,726,509 | 460 | 1.0<br>(0.1) | Whole<br>milk | 23,685,000 | 2,338 | 0.7<br>(0.0) |

<sup>1</sup> 6,772 food or beverage items were reported by participants from NHANES 2009-2016;
<sup>2</sup> The sum of the estimated weighted frequencies for all type foods or beverages reported in one day among adult participants $\geq 18$ years of NHANES 2009–2016;
<sup>3</sup> Survey weights and adjustments for the complex survey design were applied to represent the non-institutionalized U.S. population; <sup>4</sup> The frequency that a food or beverages was reported without sample weights;
<sup>5</sup> Derived from the weighted frequency of the individual food or beverage divided by the total weighted frequency of all foods and beverages (n) reported by whole sample in a single day, where n = 98,959,094 for insulin takers, n = 376,469,886 for T2D w/o taking insulin, and n = 3,245,557,852 for participants w/o diabetes. Estimated weighted percent has been abbreviated by “Wtd %”; SE stands for standard error.

**Table 3.**
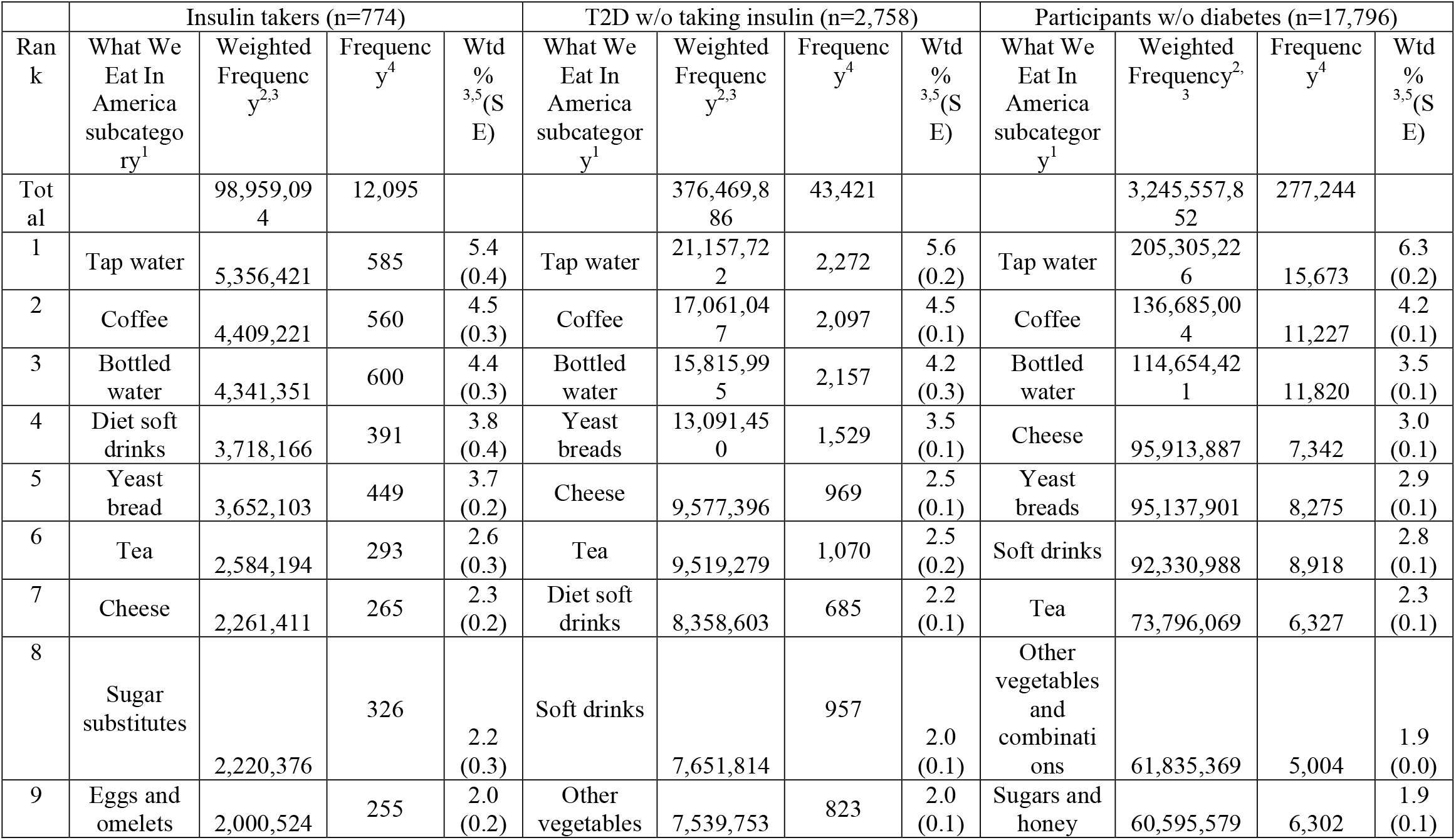
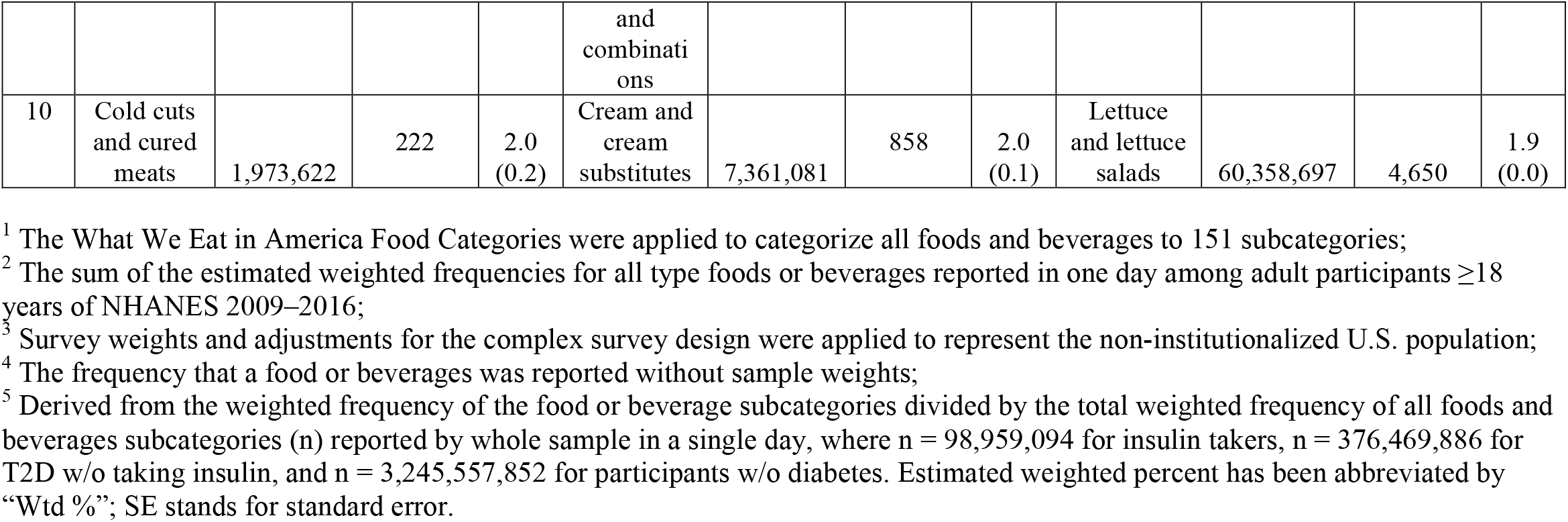
Top 10 most frequently consumed What We Eat in America foods or beverage subcategories^1^, unweighted and weighted frequency of reported foods or beverage subcategories, percent of the reported foods or beverage subcategory out of the total, and standard error of the percent of reported foods or beverage represented by the subcategory among all reported foods or beverage subcategories for U.S. adults 18 y or older taking insulin, with Type 2 diabetes but not taking insulin, and without diabetes drawn from 2009–2016 NHANES data.

|  | Insulin takers (n=774) |  |  |  | T2D w/o taking insulin (n=2,758) |  |  |  | Participants w/o diabetes (n=17,796) |  |  |  |
| --- | --- | --- | --- | --- | --- | --- | --- | --- | --- | --- | --- | --- |
| Ran<br>k | What We<br>Eat In<br>America<br>subcatego<br>ry<sup>1</sup> | Weighted<br>Frequenc<br>y<sup>2,3</sup> | Frequenc<br>y<sup>4</sup> | Wtd<br>%<br>3,5(S<br>E) | What We<br>Eat In<br>America<br>subcategor<br>y<sup>1</sup> | Weighted<br>Frequenc<br>y<sup>2,3</sup> | Frequenc<br>y<sup>4</sup> | Wtd<br>%<br>3,5(S<br>E) | What We<br>Eat In<br>America<br>subcategor<br>y<sup>1</sup> | Weighted<br>Frequency<sup>2,3</sup> | Frequenc<br>y<sup>4</sup> | Wtd<br>%<br>3,5(S<br>E) |
| Tot<br>al |  | 98,959,094 | 12,095 |  |  | 376,469,886 | 43,421 |  |  | 3,245,557,852 | 277,244 |  |
| 1 | Tap water | 5,356,421 | 585 | 5.4<br>(0.4) | Tap water | 21,157,722 | 2,272 | 5.6<br>(0.2) | Tap water | 205,305,226 | 15,673 | 6.3<br>(0.2) |
| 2 | Coffee | 4,409,221 | 560 | 4.5<br>(0.3) | Coffee | 17,061,047 | 2,097 | 4.5<br>(0.1) | Coffee | 136,685,004 | 11,227 | 4.2<br>(0.1) |
| 3 | Bottled<br>water | 4,341,351 | 600 | 4.4<br>(0.3) | Bottled<br>water | 15,815,995 | 2,157 | 4.2<br>(0.3) | Bottled<br>water | 114,654,421 | 11,820 | 3.5<br>(0.1) |
| 4 | Diet soft<br>drinks | 3,718,166 | 391 | 3.8<br>(0.4) | Yeast<br>breads | 13,091,450 | 1,529 | 3.5<br>(0.1) | Cheese | 95,913,887 | 7,342 | 3.0<br>(0.1) |
| 5 | Yeast<br>bread | 3,652,103 | 449 | 3.7<br>(0.2) | Cheese | 9,577,396 | 969 | 2.5<br>(0.1) | Yeast<br>breads | 95,137,901 | 8,275 | 2.9<br>(0.1) |
| 6 | Tea | 2,584,194 | 293 | 2.6<br>(0.3) | Tea | 9,519,279 | 1,070 | 2.5<br>(0.2) | Soft drinks | 92,330,988 | 8,918 | 2.8<br>(0.1) |
| 7 | Cheese | 2,261,411 | 265 | 2.3<br>(0.2) | Diet soft<br>drinks | 8,358,603 | 685 | 2.2<br>(0.1) | Tea | 73,796,069 | 6,327 | 2.3<br>(0.1) |
| 8 | Sugar<br>substitutes | 2,220,376 | 326 | 2.2<br>(0.3) | Soft drinks | 7,651,814 | 957 | 2.0<br>(0.1) | Other<br>vegetables<br>and<br>combinati<br>ons | 61,835,369 | 5,004 | 1.9<br>(0.0) |
| 9 | Eggs and<br>omelets | 2,000,524 | 255 | 2.0<br>(0.2) | Other<br>vegetables | 7,539,753 | 823 | 2.0<br>(0.1) | Sugars and<br>honey | 60,595,579 | 6,302 | 1.9<br>(0.1) |
|  |  |  |  |  | and<br>combinati<br>ons |  |  |  |  |  |  |  |
| 10 | Cold cuts<br>and cured<br>meats | 1,973,622 | 222 | 2.0<br>(0.2) | Cream and<br>cream<br>substitutes | 7,361,081 | 858 | 2.0<br>(0.1) | Lettuce<br>and lettuce<br>salads | 60,358,697 | 4,650 | 1.9<br>(0.0) |

<sup>1</sup> The What We Eat in America Food Categories were applied to categorize all foods and beverages to 151 subcategories;
<sup>2</sup> The sum of the estimated weighted frequencies for all type foods or beverages reported in one day among adult participants $\geq 18$ years of NHANES 2009–2016;
<sup>3</sup> Survey weights and adjustments for the complex survey design were applied to represent the non-institutionalized U.S. population;
<sup>4</sup> The frequency that a food or beverages was reported without sample weights;
<sup>5</sup> Derived from the weighted frequency of the food or beverage subcategories divided by the total weighted frequency of all foods and beverages subcategories (n) reported by whole sample in a single day, where n = 98,959,094 for insulin takers, n = 376,469,886 for T2D w/o taking insulin, and n = 3,245,557,852 for participants w/o diabetes. Estimated weighted percent has been abbreviated by “Wtd %”; SE stands for standard error.

**Table 4.**
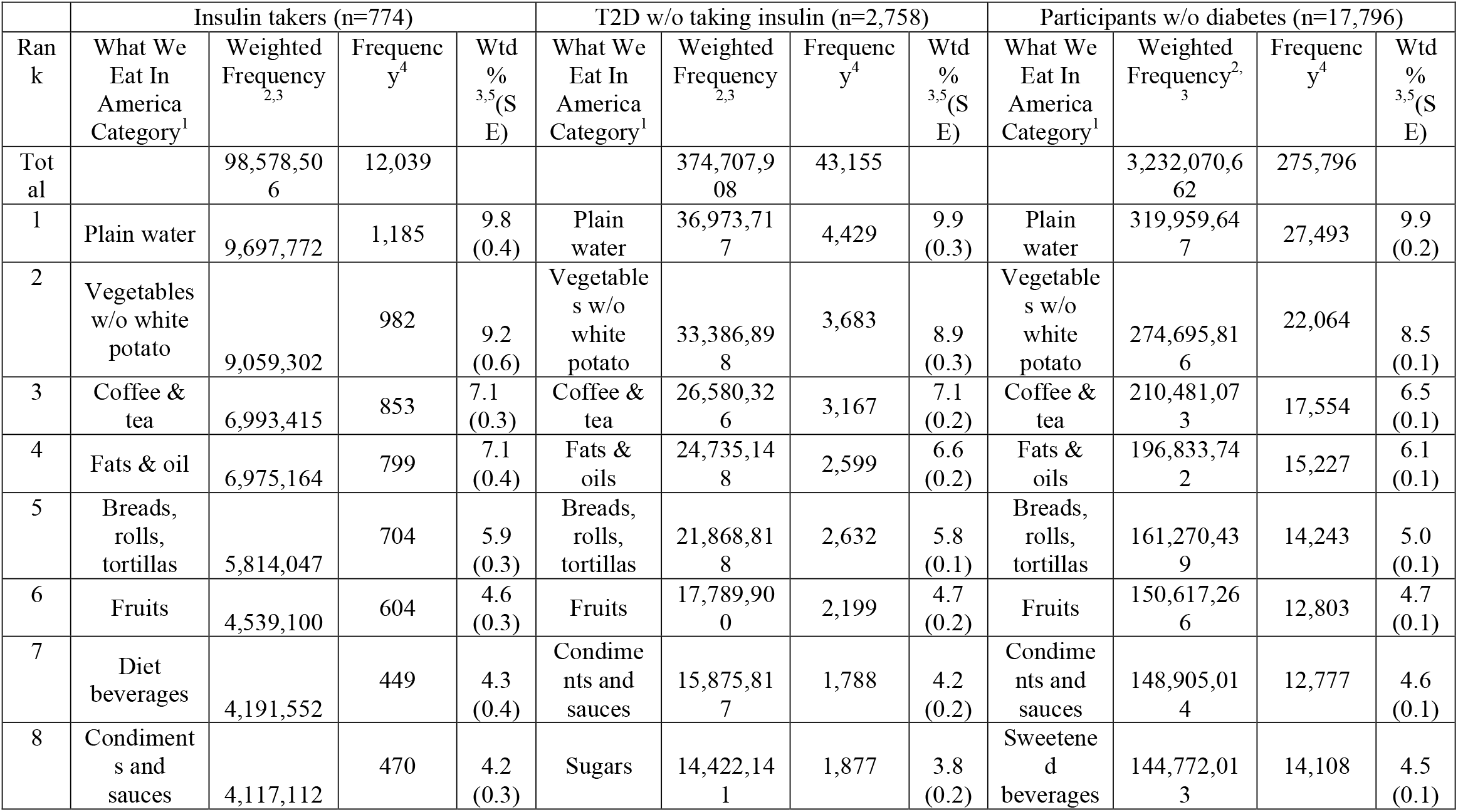
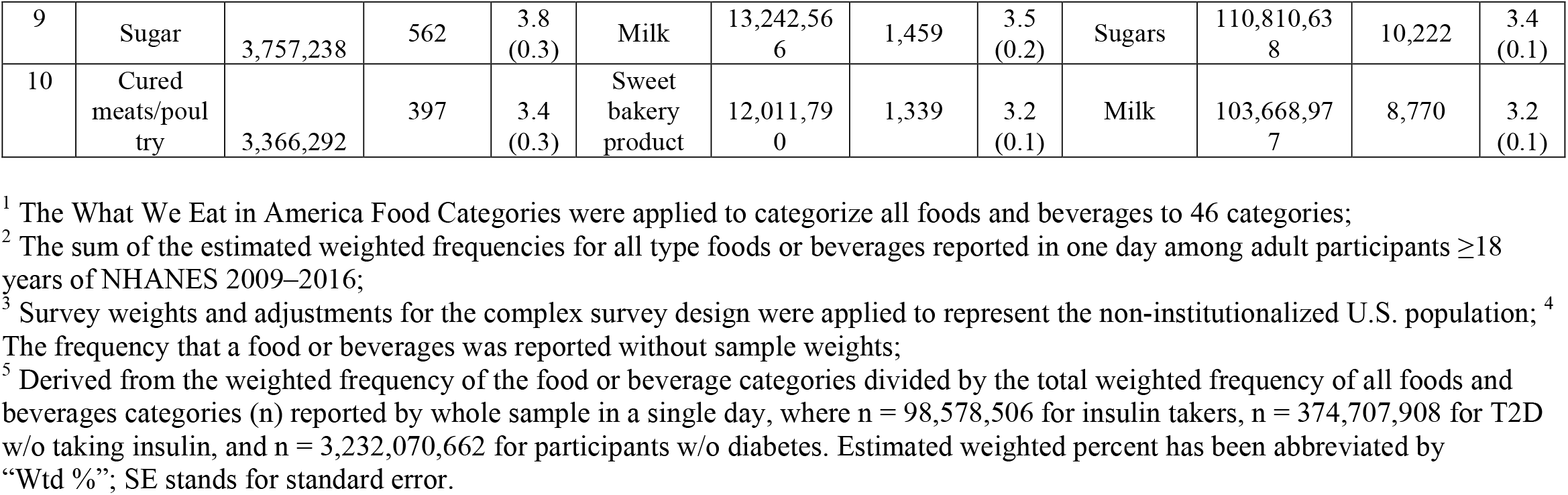
Top 10 most frequently consumed What We Eat In America food or beverage categories^1^, unweighted and weighted frequency of reported food or beverage categories, percent of the reported foods or beverage category out of total, and standard error of the percent of reported foods or beverage represented by the category among all reported foods or beverage categories for U.S. adults 18 y or older taking insulin, with Type 2 diabetes but not taking insulin, and without diabetes drawn from 2009–2016 NHANES data.

|  | Insulin takers (n=774) |  |  |  | T2D w/o taking insulin (n=2,758) |  |  |  | Participants w/o diabetes (n=17,796) |  |  |  |
| --- | --- | --- | --- | --- | --- | --- | --- | --- | --- | --- | --- | --- |
| Ran<br>k | What We<br>Eat In<br>America<br>Category<sup>1</sup> | Weighted<br>Frequency<sup>2,3</sup> | Frequenc<br>y<sup>4</sup> | Wtd<br>%<br><sup>3,5</sup>(S<br>E) | What We<br>Eat In<br>America<br>Category<sup>1</sup> | Weighted<br>Frequency<sup>2,3</sup> | Frequenc<br>y<sup>4</sup> | Wtd<br>%<br><sup>3,5</sup>(S<br>E) | What We<br>Eat In<br>America<br>Category<sup>1</sup> | Weighted<br>Frequency<sup>2,3</sup> | Frequenc<br>y<sup>4</sup> | Wtd<br>%<br><sup>3,5</sup>(S<br>E) |
| Tot<br>al |  | 98,578,506 | 12,039 |  |  | 374,707,908 | 43,155 |  |  | 3,232,070,662 | 275,796 |  |
| 1 | Plain water | 9,697,772 | 1,185 | 9.8<br>(0.4) | Plain water | 36,973,717 | 4,429 | 9.9<br>(0.3) | Plain water | 319,959,647 | 27,493 | 9.9<br>(0.2) |
| 2 | Vegetables w/o white potato | 9,059,302 | 982 | 9.2<br>(0.6) | Vegetables w/o white potato | 33,386,898 | 3,683 | 8.9<br>(0.3) | Vegetables w/o white potato | 274,695,816 | 22,064 | 8.5<br>(0.1) |
| 3 | Coffee & tea | 6,993,415 | 853 | 7.1<br>(0.3) | Coffee & tea | 26,580,326 | 3,167 | 7.1<br>(0.2) | Coffee & tea | 210,481,073 | 17,554 | 6.5<br>(0.1) |
| 4 | Fats & oil | 6,975,164 | 799 | 7.1<br>(0.4) | Fats & oils | 24,735,148 | 2,599 | 6.6<br>(0.2) | Fats & oils | 196,833,742 | 15,227 | 6.1<br>(0.1) |
| 5 | Breads, rolls, tortillas | 5,814,047 | 704 | 5.9<br>(0.3) | Breads, rolls, tortillas | 21,868,818 | 2,632 | 5.8<br>(0.1) | Breads, rolls, tortillas | 161,270,439 | 14,243 | 5.0<br>(0.1) |
| 6 | Fruits | 4,539,100 | 604 | 4.6<br>(0.3) | Fruits | 17,789,900 | 2,199 | 4.7<br>(0.2) | Fruits | 150,617,266 | 12,803 | 4.7<br>(0.1) |
| 7 | Diet beverages | 4,191,552 | 449 | 4.3<br>(0.4) | Condiments and sauces | 15,875,817 | 1,788 | 4.2<br>(0.2) | Condiments and sauces | 148,905,014 | 12,777 | 4.6<br>(0.1) |
| 8 | Condiments and sauces | 4,117,112 | 470 | 4.2<br>(0.3) | Sugars | 14,422,141 | 1,877 | 3.8<br>(0.2) | Sweetened beverages | 144,772,013 | 14,108 | 4.5<br>(0.1) |
| 9 | Sugar | 3,757,238 | 562 | 3.8<br>(0.3) | Milk | 13,242,566 | 1,459 | 3.5<br>(0.2) | Sugars | 110,810,638 | 10,222 | 3.4<br>(0.1) |
| 10 | Cured meats/poultry | 3,366,292 | 397 | 3.4<br>(0.3) | Sweet bakery product | 12,011,790 | 1,339 | 3.2<br>(0.1) | Milk | 103,668,977 | 8,770 | 3.2<br>(0.1) |

<sup>1</sup> The What We Eat in America Food Categories were applied to categorize all foods and beverages to 46 categories;
<sup>2</sup> The sum of the estimated weighted frequencies for all type foods or beverages reported in one day among adult participants $\geq 18$ years of NHANES 2009–2016;
<sup>3</sup> Survey weights and adjustments for the complex survey design were applied to represent the non-institutionalized U.S. population; <sup>4</sup> The frequency that a food or beverages was reported without sample weights;
<sup>5</sup> Derived from the weighted frequency of the food or beverage categories divided by the total weighted frequency of all foods and beverages categories (n) reported by whole sample in a single day, where n = 98,578,506 for insulin takers, n = 374,707,908 for T2D w/o taking insulin, and n = 3,232,070,662 for participants w/o diabetes. Estimated weighted percent has been abbreviated by “Wtd %”; SE stands for standard error.

### 3.3 The highest energy contributing food items, food subcategories, and food categories

The top 10 highest energy contributing food items, food subcategories, and food categories for insulin takers, T2D not using insulin, and participants without diabetes are shown in **Tables 5-7**. Foods are presented in descending weighted order according to their energy contribution by their FNDDS short food descriptions. In energy contribution lists, 5 food items (Table 5), 6 food subcategories (Table 6), and 5 food categories (Table 7) were similar among the three groups but their rankings were different. Regular soft drinks did not rank in the top 10 most energy contributing food subcategories list among insulin takers, but was ranked 5^th^ among those with T2D who did not use insulin, and 2^nd^ among participants without diabetes (Table 6). Furthermore, insulin takers consumed less energy from food subcategories with added sugars compared with the other two groups (cakes and pies were 6^th^ and cookies and brownies 7^th^ among insulin takers; cakes and pies were 3^rd^ and cookies and brownies 6^th^ among those with T2D not taking insulin; cookies and brownies were 8^th^ and cakes and pies 9^th^ among participants without diabetes). More protein food subcategories ranked high within the top 10 list (whole pieces of chicken ranked 3^rd^ place, eggs and omelets ranked 4^th^ place, meat mixed dishes ranked 5^th^ place, cold cuts and cured meats ranked 10^th^ place) among insulin takers compared to the other two groups (whole pieces of chicken ranked 4^th^ place, eggs and omelets 9^th^ place, meat mixed dishes 10^th^ place in those with T2D not using insulin, and whole pieces of chicken ranked 5^th^ place in participants without diabetes) (Table 6).

**Table 5.**
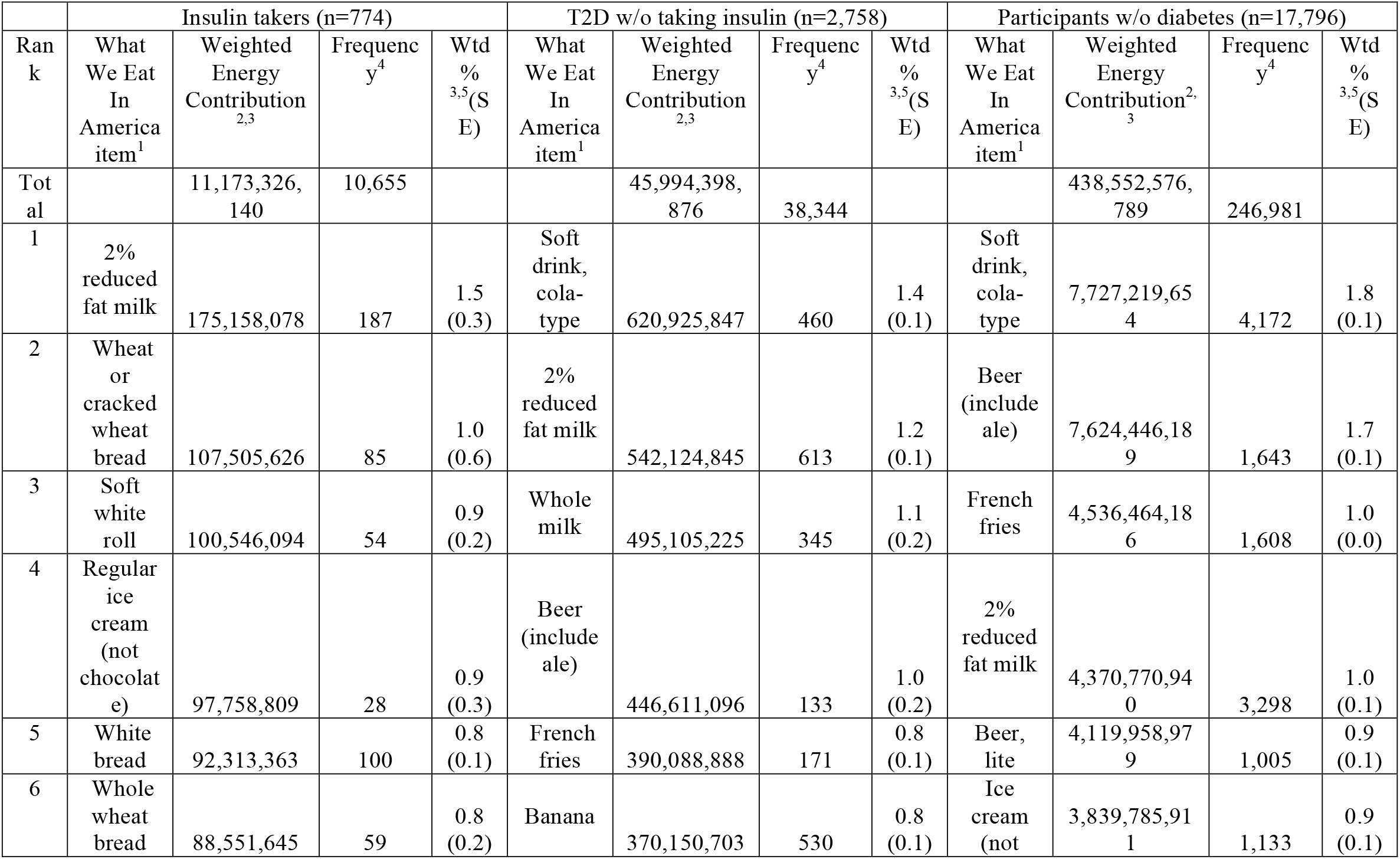
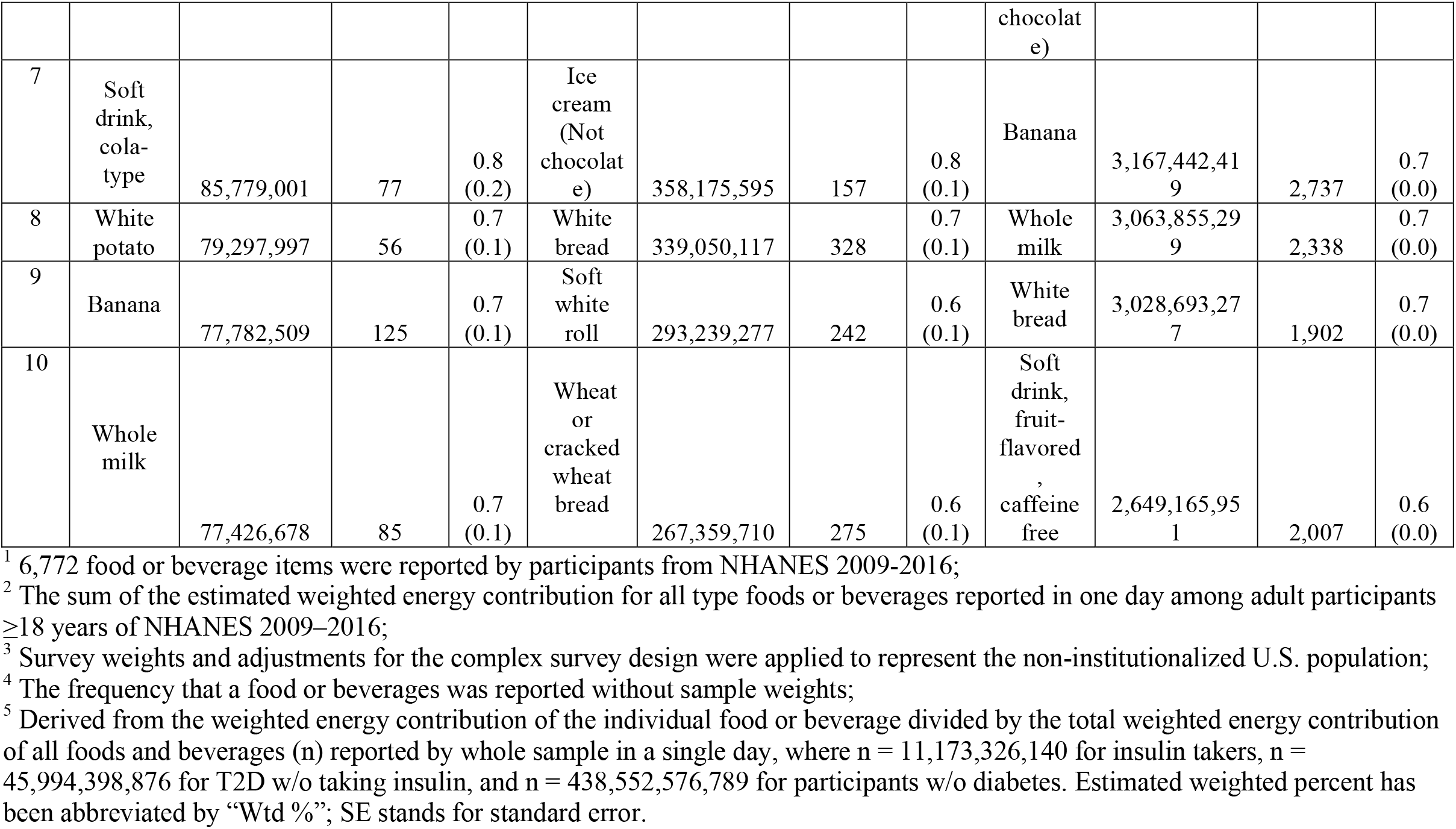
Top 10 foods items or beverages contributing the most to total unweighted and weighted energy, percent of the total represented by the reported food or beverage item^1^, and standard error of the percent of the total represented by the reported food or beverage item among all reported foods or beverage items for U.S. adults 18 y or older taking insulin, with Type 2 diabetes but not taking insulin, and without diabetes drawn from 2009–2016 NHANES data.

|  | Insulin takers (n=774) |  |  |  | T2D w/o taking insulin (n=2,758) |  |  |  | Participants w/o diabetes (n=17,796) |  |  |  |
| --- | --- | --- | --- | --- | --- | --- | --- | --- | --- | --- | --- | --- |
| Ran<br>k | What<br>We Eat<br>In<br>America<br>item<sup>1</sup> | Weighted<br>Energy<br>Contribution<sup>2,3</sup> | Frequenc<br>y<sup>4</sup> | Wtd<br>%<br><sup>3,5</sup>(S<br>E) | What<br>We Eat<br>In<br>America<br>item<sup>1</sup> | Weighted<br>Energy<br>Contribution<sup>2,3</sup> | Frequenc<br>y<sup>4</sup> | Wtd<br>%<br><sup>3,5</sup>(S<br>E) | What<br>We Eat<br>In<br>America<br>item<sup>1</sup> | Weighted<br>Energy<br>Contribution<sup>2,3</sup> | Frequenc<br>y<sup>4</sup> | Wtd<br>%<br><sup>3,5</sup>(S<br>E) |
| Tot<br>al |  | 11,173,326,<br>140 | 10,655 |  |  | 45,994,398,<br>876 | 38,344 |  |  | 438,552,576,<br>789 | 246,981 |  |
| 1 | 2%<br>reduced<br>fat milk | 175,158,078 | 187 | 1.5<br>(0.3) | Soft<br>drink,<br>cola-type | 620,925,847 | 460 | 1.4<br>(0.1) | Soft<br>drink,<br>cola-type | 7,727,219,65<br>4 | 4,172 | 1.8<br>(0.1) |
| 2 | Wheat<br>or<br>cracked<br>wheat<br>bread | 107,505,626 | 85 | 1.0<br>(0.6) | 2%<br>reduced<br>fat milk | 542,124,845 | 613 | 1.2<br>(0.1) | Beer<br>(include<br>ale) | 7,624,446,18<br>9 | 1,643 | 1.7<br>(0.1) |
| 3 | Soft<br>white<br>roll | 100,546,094 | 54 | 0.9<br>(0.2) | Whole<br>milk | 495,105,225 | 345 | 1.1<br>(0.2) | French<br>fries | 4,536,464,18<br>6 | 1,608 | 1.0<br>(0.0) |
| 4 | Regular<br>ice<br>cream<br>(not<br>chocolat<br>e) | 97,758,809 | 28 | 0.9<br>(0.3) | Beer<br>(include<br>ale) | 446,611,096 | 133 | 1.0<br>(0.2) | 2%<br>reduced<br>fat milk | 4,370,770,94<br>0 | 3,298 | 1.0<br>(0.1) |
| 5 | White<br>bread | 92,313,363 | 100 | 0.8<br>(0.1) | French<br>fries | 390,088,888 | 171 | 0.8<br>(0.1) | Beer,<br>lite | 4,119,958,97<br>9 | 1,005 | 0.9<br>(0.1) |
| 6 | Whole<br>wheat<br>bread | 88,551,645 | 59 | 0.8<br>(0.2) | Banana | 370,150,703 | 530 | 0.8<br>(0.1) | Ice<br>cream<br>(not | 3,839,785,91<br>1 | 1,133 | 0.9<br>(0.1) |
|  |  |  |  |  |  |  |  |  | chocolate) |  |  |  |
| 7 | Soft drink, cola-type | 85,779,001 | 77 | 0.8<br>(0.2) | Ice cream (Not chocolate) | 358,175,595 | 157 | 0.8<br>(0.1) | Banana | 3,167,442,419 | 2,737 | 0.7<br>(0.0) |
| 8 | White potato | 79,297,997 | 56 | 0.7<br>(0.1) | White bread | 339,050,117 | 328 | 0.7<br>(0.1) | Whole milk | 3,063,855,299 | 2,338 | 0.7<br>(0.0) |
| 9 | Banana | 77,782,509 | 125 | 0.7<br>(0.1) | Soft white roll | 293,239,277 | 242 | 0.6<br>(0.1) | White bread | 3,028,693,277 | 1,902 | 0.7<br>(0.0) |
| 10 | Whole milk | 77,426,678 | 85 | 0.7<br>(0.1) | Wheat or cracked wheat bread | 267,359,710 | 275 | 0.6<br>(0.1) | Soft drink, fruit-flavored, caffeine free | 2,649,165,951 | 2,007 | 0.6<br>(0.0) |

<sup>1</sup> 6,772 food or beverage items were reported by participants from NHANES 2009-2016;
<sup>2</sup> The sum of the estimated weighted energy contribution for all type foods or beverages reported in one day among adult participants $\geq 18$ years of NHANES 2009–2016;
<sup>3</sup> Survey weights and adjustments for the complex survey design were applied to represent the non-institutionalized U.S. population;
<sup>4</sup> The frequency that a food or beverages was reported without sample weights;
<sup>5</sup> Derived from the weighted energy contribution of the individual food or beverage divided by the total weighted energy contribution of all foods and beverages (n) reported by whole sample in a single day, where n = 11,173,326,140 for insulin takers, n = 45,994,398,876 for T2D w/o taking insulin, and n = 438,552,576,789 for participants w/o diabetes. Estimated weighted percent has been abbreviated by “Wtd %”; SE stands for standard error.

**Table 6.**
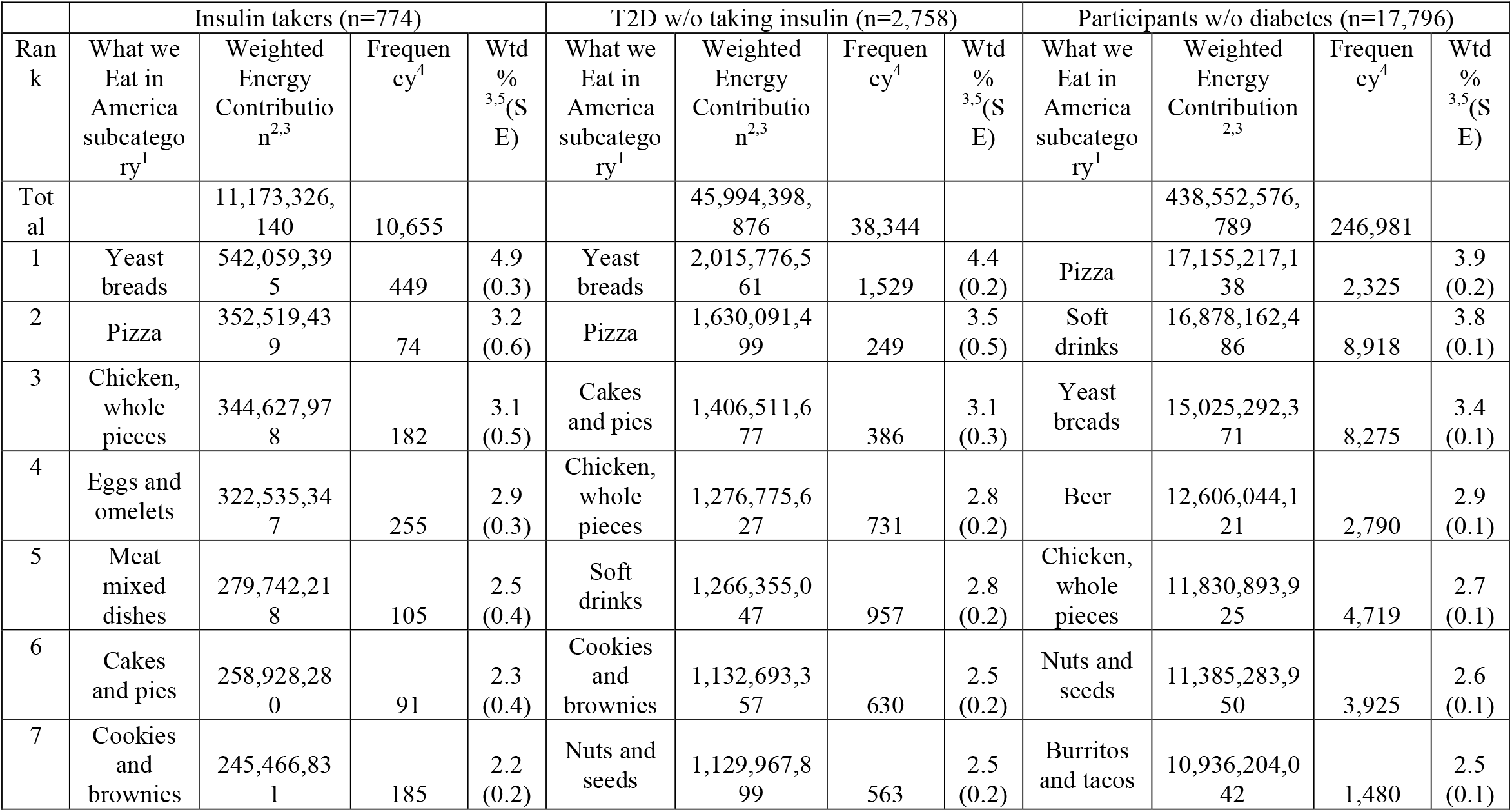
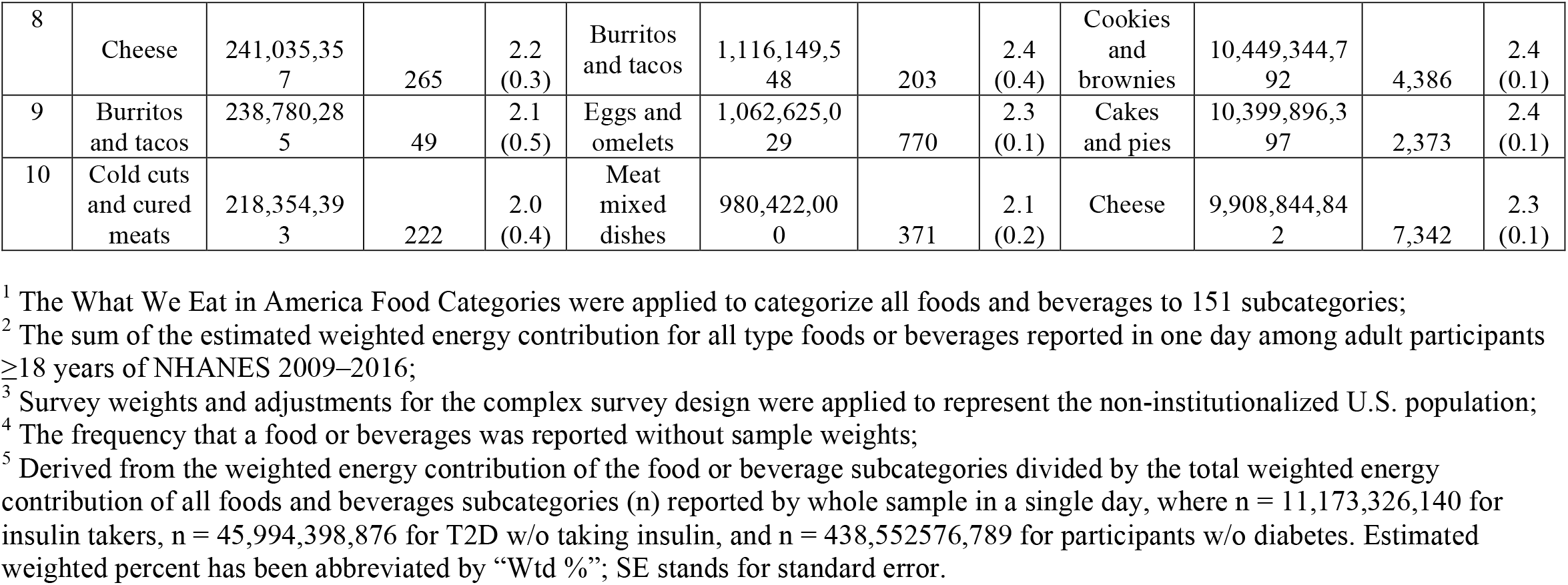
Top 10 highest energy contributing What We Eat in America food and beverage subcategories^1^ contributing the most to unweighted and weighted energy, percent of the total represented by the reported food or beverage subcategory, and standard error of the percent of the total represented by the reported food or beverage subcategory among all reported food or beverage subcategories for U.S. adults 18 y or older taking insulin, with Type 2 diabetes but not taking insulin, and without diabetes drawn from 2009–2016 NHANES data.

|  | Insulin takers (n=774) |  |  |  | T2D w/o taking insulin (n=2,758) |  |  |  | Participants w/o diabetes (n=17,796) |  |  |  |
| --- | --- | --- | --- | --- | --- | --- | --- | --- | --- | --- | --- | --- |
| Ran<br>k | What we<br>Eat in<br>America<br>subcatego<br>ry<sup>1</sup> | Weighted<br>Energy<br>Contributio<br>n<sup>2,3</sup> | Frequen<br>cy<sup>4</sup> | Wtd<br>%<br><sup>3,5</sup>(S<br>E) | What we<br>Eat in<br>America<br>subcatego<br>ry<sup>1</sup> | Weighted<br>Energy<br>Contributio<br>n<sup>2,3</sup> | Frequen<br>cy<sup>4</sup> | Wtd<br>%<br><sup>3,5</sup>(S<br>E) | What we<br>Eat in<br>America<br>subcatego<br>ry<sup>1</sup> | Weighted<br>Energy<br>Contribution<br><sup>2,3</sup> | Frequen<br>cy<sup>4</sup> | Wtd<br>%<br><sup>3,5</sup>(S<br>E) |
| Tot<br>al |  | 11,173,326,<br>140 | 10,655 |  |  | 45,994,398,<br>876 | 38,344 |  |  | 438,552,576,<br>789 | 246,981 |  |
| 1 | Yeast<br>breads | 542,059,39<br>5 | 449 | 4.9<br>(0.3) | Yeast<br>breads | 2,015,776,5<br>61 | 1,529 | 4.4<br>(0.2) | Pizza | 17,155,217,1<br>38 | 2,325 | 3.9<br>(0.2) |
| 2 | Pizza | 352,519,43<br>9 | 74 | 3.2<br>(0.6) | Pizza | 1,630,091,4<br>99 | 249 | 3.5<br>(0.5) | Soft<br>drinks | 16,878,162,4<br>86 | 8,918 | 3.8<br>(0.1) |
| 3 | Chicken,<br>whole<br>pieces | 344,627,97<br>8 | 182 | 3.1<br>(0.5) | Cakes<br>and pies | 1,406,511,6<br>77 | 386 | 3.1<br>(0.3) | Yeast<br>breads | 15,025,292,3<br>71 | 8,275 | 3.4<br>(0.1) |
| 4 | Eggs and<br>omelets | 322,535,34<br>7 | 255 | 2.9<br>(0.3) | Chicken,<br>whole<br>pieces | 1,276,775,6<br>27 | 731 | 2.8<br>(0.2) | Beer | 12,606,044,1<br>21 | 2,790 | 2.9<br>(0.1) |
| 5 | Meat<br>mixed<br>dishes | 279,742,21<br>8 | 105 | 2.5<br>(0.4) | Soft<br>drinks | 1,266,355,0<br>47 | 957 | 2.8<br>(0.2) | Chicken,<br>whole<br>pieces | 11,830,893,9<br>25 | 4,719 | 2.7<br>(0.1) |
| 6 | Cakes<br>and pies | 258,928,28<br>0 | 91 | 2.3<br>(0.4) | Cookies<br>and<br>brownies | 1,132,693,3<br>57 | 630 | 2.5<br>(0.2) | Nuts and<br>seeds | 11,385,283,9<br>50 | 3,925 | 2.6<br>(0.1) |
| 7 | Cookies<br>and<br>brownies | 245,466,83<br>1 | 185 | 2.2<br>(0.2) | Nuts and<br>seeds | 1,129,967,8<br>99 | 563 | 2.5<br>(0.2) | Burritos<br>and tacos | 10,936,204,0<br>42 | 1,480 | 2.5<br>(0.1) |
| 8 | Cheese | 241,035,357 | 265 | 2.2<br>(0.3) | Burritos and tacos | 1,116,149,548 | 203 | 2.4<br>(0.4) | Cookies and brownies | 10,449,344,792 | 4,386 | 2.4<br>(0.1) |
| 9 | Burritos and tacos | 238,780,285 | 49 | 2.1<br>(0.5) | Eggs and omelets | 1,062,625,029 | 770 | 2.3<br>(0.1) | Cakes and pies | 10,399,896,397 | 2,373 | 2.4<br>(0.1) |
| 10 | Cold cuts and cured meats | 218,354,393 | 222 | 2.0<br>(0.4) | Meat mixed dishes | 980,422,000 | 371 | 2.1<br>(0.2) | Cheese | 9,908,844,842 | 7,342 | 2.3<br>(0.1) |

<sup>1</sup> The What We Eat in America Food Categories were applied to categorize all foods and beverages to 151 subcategories;
<sup>2</sup> The sum of the estimated weighted energy contribution for all type foods or beverages reported in one day among adult participants $\geq 18$ years of NHANES 2009–2016;
<sup>3</sup> Survey weights and adjustments for the complex survey design were applied to represent the non-institutionalized U.S. population;
<sup>4</sup> The frequency that a food or beverages was reported without sample weights;
<sup>5</sup> Derived from the weighted energy contribution of the food or beverage subcategories divided by the total weighted energy contribution of all foods and beverages subcategories (n) reported by whole sample in a single day, where n = 11,173,326,140 for insulin takers, n = 45,994,398,876 for T2D w/o taking insulin, and n = 438,552,576,789 for participants w/o diabetes. Estimated weighted percent has been abbreviated by “Wtd %”; SE stands for standard error.

**Table 7.**
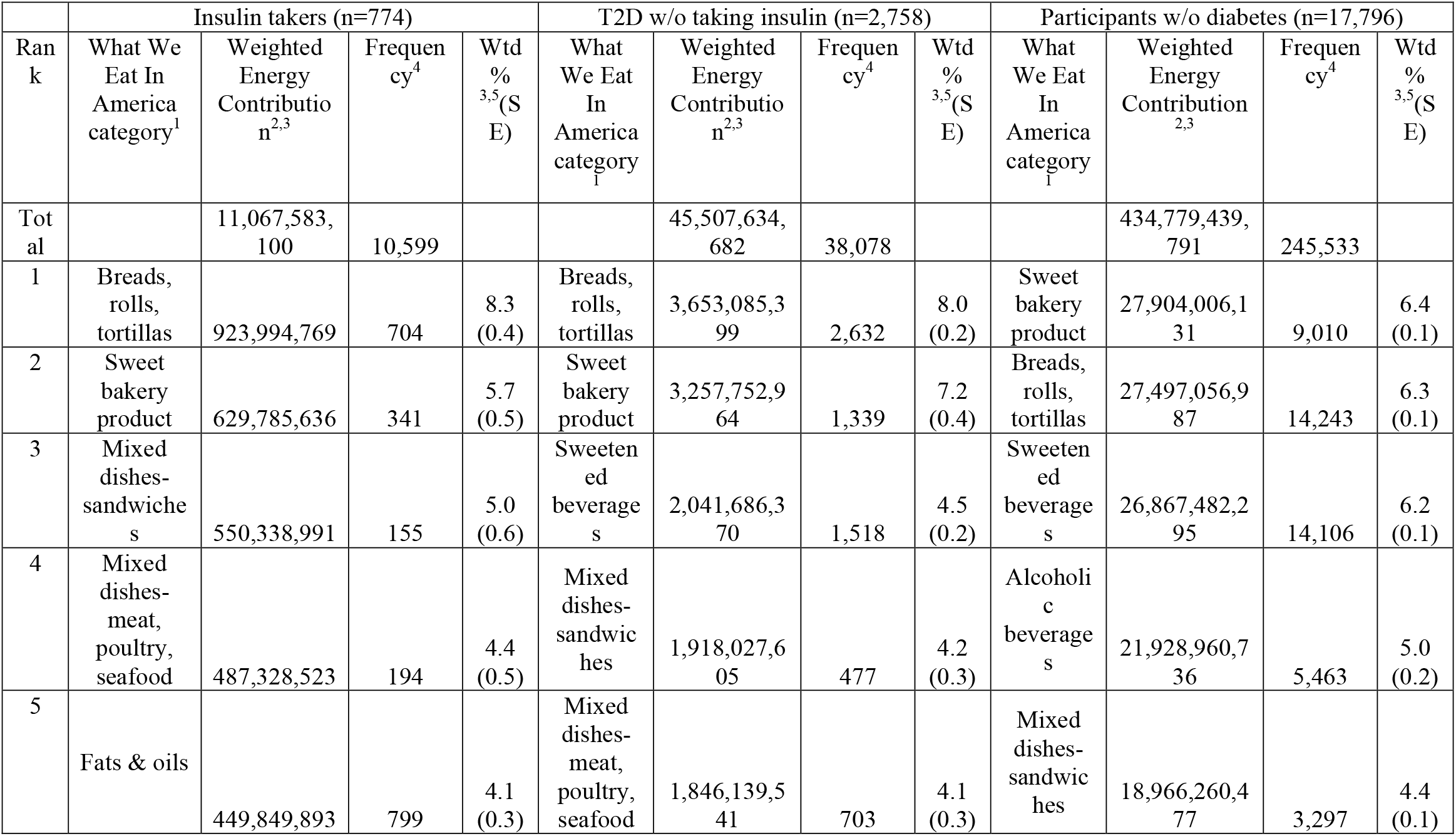
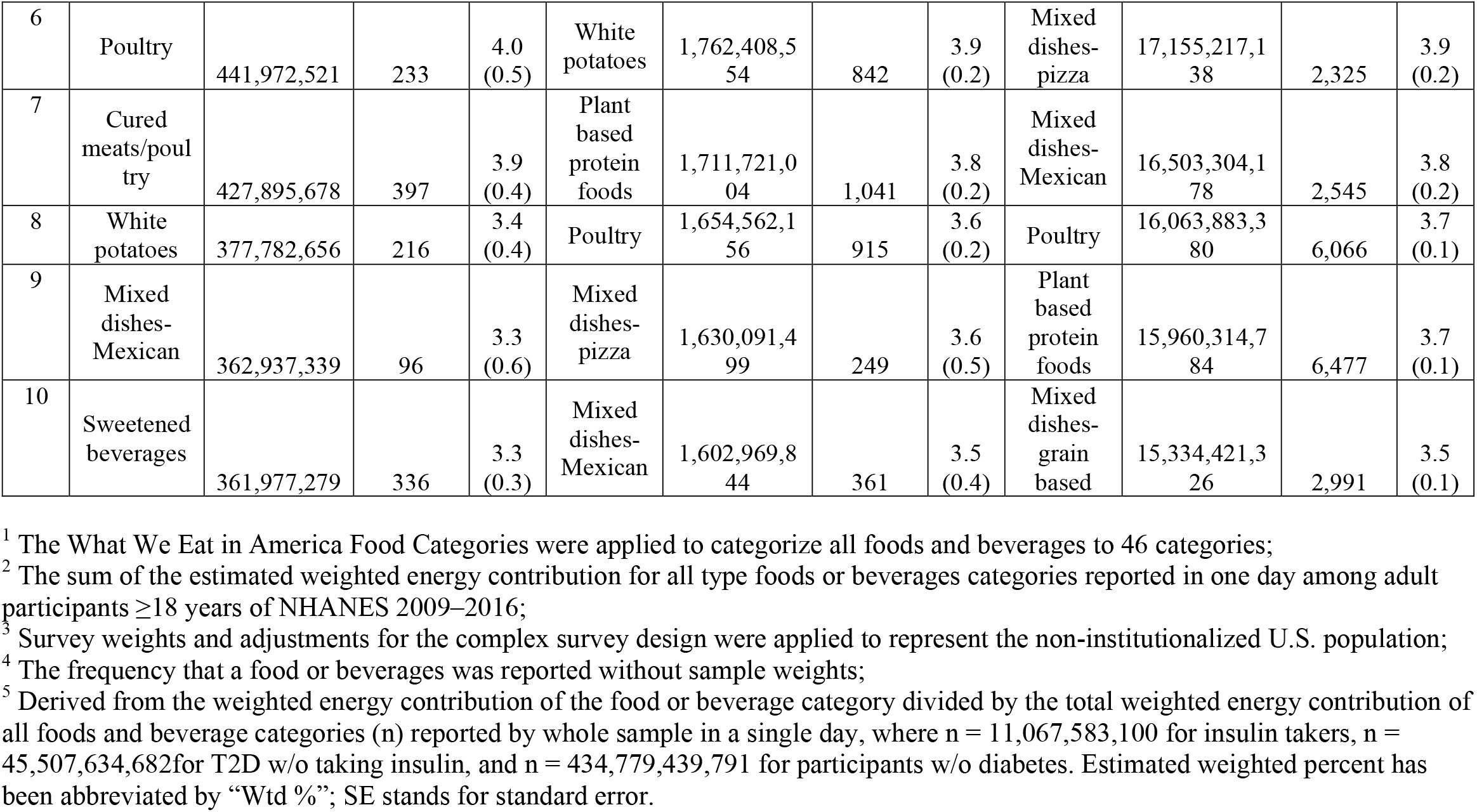
Top 10 highest energy contributing What We Eat in America food and beverage categories^1^ contributing the most to unweighted and weighted energy, percent of the total represented by the reported food or beverage category, and standard error of the percent of the total represented by the reported food or beverage category among all reported food or beverage categories for U.S. adults 18 y or older taking insulin, with Type 2 diabetes but not taking insulin, and without diabetes drawn from 2009–2016 NHANES data.

|  | Insulin takers (n=774) |  |  |  | T2D w/o taking insulin (n=2,758) |  |  |  | Participants w/o diabetes (n=17,796) |  |  |  |
| --- | --- | --- | --- | --- | --- | --- | --- | --- | --- | --- | --- | --- |
| Ran<br>k | What We<br>Eat In<br>America<br>category<sup>1</sup> | Weighted<br>Energy<br>Contributio<br>n<sup>2,3</sup> | Frequen<br>cy<sup>4</sup> | Wtd<br>%<br><sup>3,5</sup>(S<br>E) | What<br>We Eat<br>In<br>America<br>category<sup>1</sup> | Weighted<br>Energy<br>Contributio<br>n<sup>2,3</sup> | Frequen<br>cy<sup>4</sup> | Wtd<br>%<br><sup>3,5</sup>(S<br>E) | What<br>We Eat<br>In<br>America<br>category<sup>1</sup> | Weighted<br>Energy<br>Contribution<br><sup>2,3</sup> | Frequen<br>cy<sup>4</sup> | Wtd<br>%<br><sup>3,5</sup>(S<br>E) |
| Tot<br>al |  | 11,067,583,<br>100 | 10,599 |  |  | 45,507,634,<br>682 | 38,078 |  |  | 434,779,439,<br>791 | 245,533 |  |
| 1 | Breads,<br>rolls,<br>tortillas | 923,994,769 | 704 | 8.3<br>(0.4) | Breads,<br>rolls,<br>tortillas | 3,653,085,3<br>99 | 2,632 | 8.0<br>(0.2) | Sweet<br>bakery<br>product | 27,904,006,1<br>31 | 9,010 | 6.4<br>(0.1) |
| 2 | Sweet<br>bakery<br>product | 629,785,636 | 341 | 5.7<br>(0.5) | Sweet<br>bakery<br>product | 3,257,752,9<br>64 | 1,339 | 7.2<br>(0.4) | Breads,<br>rolls,<br>tortillas | 27,497,056,9<br>87 | 14,243 | 6.3<br>(0.1) |
| 3 | Mixed<br>dishes-<br>sandwiche<br>s | 550,338,991 | 155 | 5.0<br>(0.6) | Sweeten<br>ed<br>beverage<br>s | 2,041,686,3<br>70 | 1,518 | 4.5<br>(0.2) | Sweeten<br>ed<br>beverage<br>s | 26,867,482,2<br>95 | 14,106 | 6.2<br>(0.1) |
| 4 | Mixed<br>dishes-<br>meat,<br>poultry,<br>seafood | 487,328,523 | 194 | 4.4<br>(0.5) | Mixed<br>dishes-<br>sandwic<br>hes | 1,918,027,6<br>05 | 477 | 4.2<br>(0.3) | Alcoholi<br>c<br>beverage<br>s | 21,928,960,7<br>36 | 5,463 | 5.0<br>(0.2) |
| 5 | Fats & oils | 449,849,893 | 799 | 4.1<br>(0.3) | Mixed<br>dishes-<br>meat,<br>poultry,<br>seafood | 1,846,139,5<br>41 | 703 | 4.1<br>(0.3) | Mixed<br>dishes-<br>sandwic<br>hes | 18,966,260,4<br>77 | 3,297 | 4.4<br>(0.1) |
| 6 | Poultry | 441,972,521 | 233 | 4.0<br>(0.5) | White<br>potatoes | 1,762,408,5<br>54 | 842 | 3.9<br>(0.2) | Mixed<br>dishes-<br>pizza | 17,155,217,1<br>38 | 2,325 | 3.9<br>(0.2) |
| 7 | Cured<br>meats/poul<br>try | 427,895,678 | 397 | 3.9<br>(0.4) | Plant<br>based<br>protein<br>foods | 1,711,721,0<br>04 | 1,041 | 3.8<br>(0.2) | Mixed<br>dishes-Mexican | 16,503,304,1<br>78 | 2,545 | 3.8<br>(0.2) |
| 8 | White<br>potatoes | 377,782,656 | 216 | 3.4<br>(0.4) | Poultry | 1,654,562,1<br>56 | 915 | 3.6<br>(0.2) | Poultry | 16,063,883,3<br>80 | 6,066 | 3.7<br>(0.1) |
| 9 | Mixed<br>dishes-Mexican | 362,937,339 | 96 | 3.3<br>(0.6) | Mixed<br>dishes-<br>pizza | 1,630,091,4<br>99 | 249 | 3.6<br>(0.5) | Plant<br>based<br>protein<br>foods | 15,960,314,7<br>84 | 6,477 | 3.7<br>(0.1) |
| 10 | Sweetened<br>beverages | 361,977,279 | 336 | 3.3<br>(0.3) | Mixed<br>dishes-Mexican | 1,602,969,8<br>44 | 361 | 3.5<br>(0.4) | Mixed<br>dishes-<br>grain<br>based | 15,334,421,3<br>26 | 2,991 | 3.5<br>(0.1) |

<sup>1</sup> The What We Eat in America Food Categories were applied to categorize all foods and beverages to 46 categories;
<sup>2</sup> The sum of the estimated weighted energy contribution for all type foods or beverages categories reported in one day among adult participants $\geq 18$ years of NHANES 2009–2016;
<sup>3</sup> Survey weights and adjustments for the complex survey design were applied to represent the non-institutionalized U.S. population;
<sup>4</sup> The frequency that a food or beverages was reported without sample weights;
<sup>5</sup> Derived from the weighted energy contribution of the food or beverage category divided by the total weighted energy contribution of all foods and beverage categories (n) reported by whole sample in a single day, where n = 11,067,583,100 for insulin takers, n = 45,507,634,682 for T2D w/o taking insulin, and n = 434,779,439,791 for participants w/o diabetes. Estimated weighted percent has been abbreviated by “Wtd %”; SE stands for standard error.

### 3.4 Broad Food Categories Intake by Frequency and Energy Contribution

The results of the comparison of the frequency and energy contribution of the broad WWEIA food or beverage categories among the three groups are shown in **Table 8**. Broad categories of grains and alcohol were statistically significantly different in both intake frequency and energy contribution across the three groups. From insulin takers to T2D not using insulin to participants without diabetes, grains exhibited a decreasing trend in consumption frequency and energy contribution, while alcohol showed an increasing trend. Snacks / sweets were statistically significantly different in consumption frequency among the three groups, which showed an increasing trend from insulin takers to T2D not using insulin to participants without diabetes. Protein foods, vegetables, beverages, water, and other were statistically significantly different in energy contribution across the three groups. From insulin takers to participants without diabetes, protein foods and vegetables exhibited a decreasing trend in energy contribution, while beverages, water, and other showed an increasing trend.

**Table 8.**
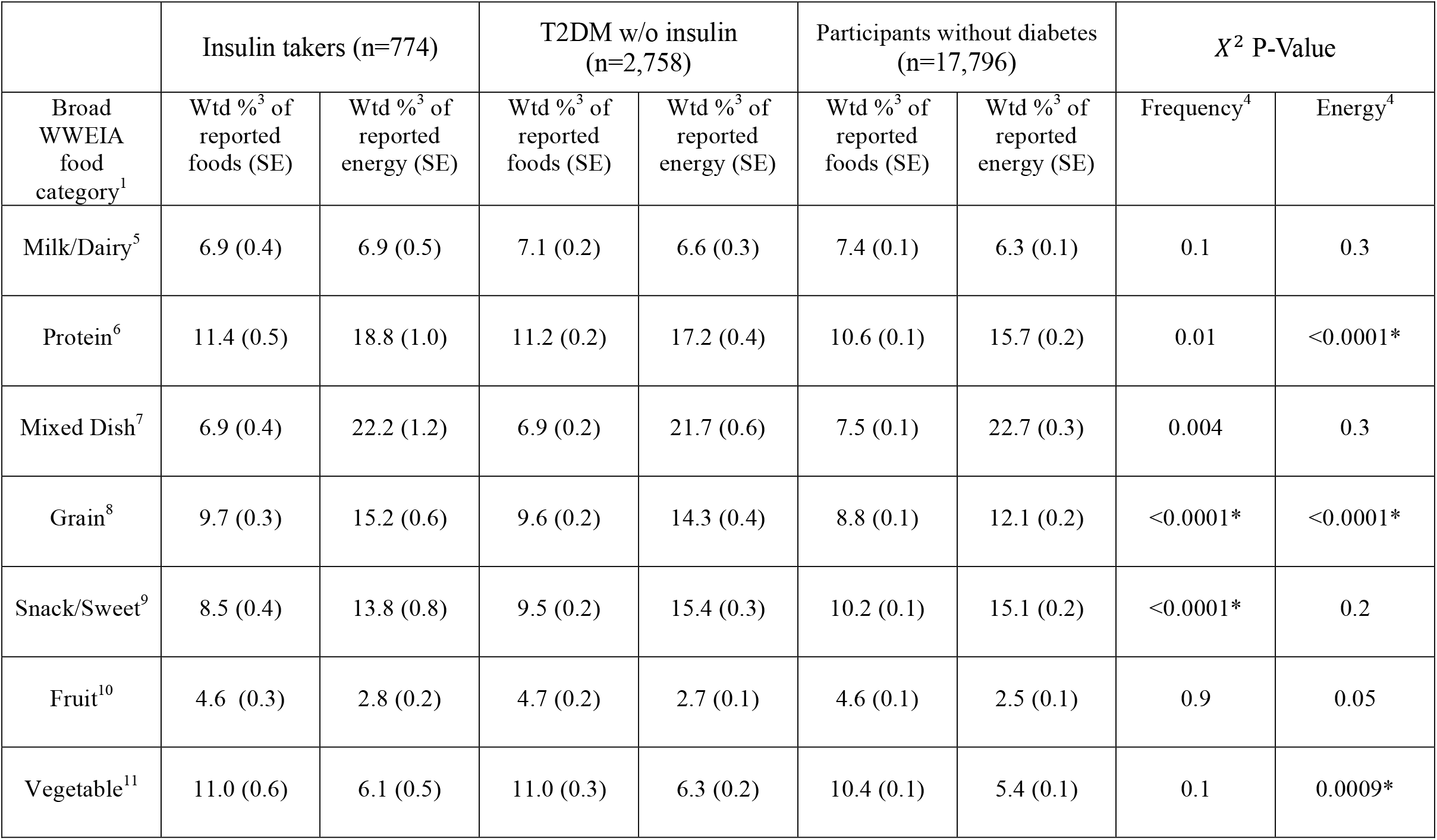
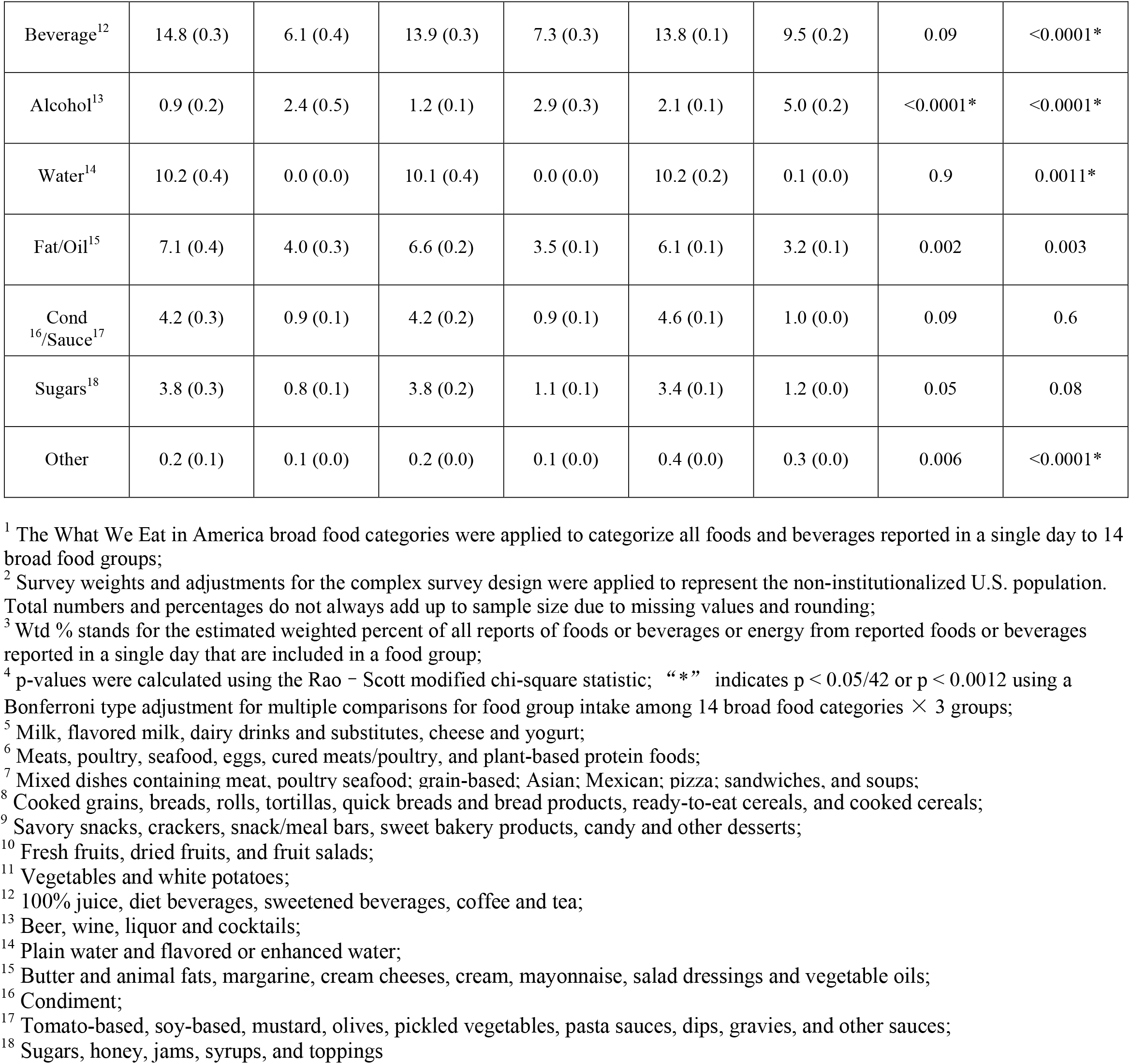
Broad What We Eat In America food and beverage category^1^ intake comparisons by frequency and energy among U.S. adults 18 y or older taking insulin, with Type 2 diabetes but not taking insulin, and without diabetes drawn from NHANES 2009–2016^2^

|  | Insulin takers (n=774) |  | T2DM w/o insulin (n=2,758) |  | Participants without diabetes (n=17,796) |  | X<sup>2</sup> P-Value |  |
| --- | --- | --- | --- | --- | --- | --- | --- | --- |
| Broad WWEIA food category<sup>1</sup> | Wtd %<sup>3</sup> of reported foods (SE) | Wtd %<sup>3</sup> of reported energy (SE) | Wtd %<sup>3</sup> of reported foods (SE) | Wtd %<sup>3</sup> of reported energy (SE) | Wtd %<sup>3</sup> of reported foods (SE) | Wtd %<sup>3</sup> of reported energy (SE) | Frequency<sup>4</sup> | Energy<sup>4</sup> |
| Milk/Dairy<sup>5</sup> | 6.9 (0.4) | 6.9 (0.5) | 7.1 (0.2) | 6.6 (0.3) | 7.4 (0.1) | 6.3 (0.1) | 0.1 | 0.3 |
| Protein<sup>6</sup> | 11.4 (0.5) | 18.8 (1.0) | 11.2 (0.2) | 17.2 (0.4) | 10.6 (0.1) | 15.7 (0.2) | 0.01 | <0.0001* |
| Mixed Dish<sup>7</sup> | 6.9 (0.4) | 22.2 (1.2) | 6.9 (0.2) | 21.7 (0.6) | 7.5 (0.1) | 22.7 (0.3) | 0.004 | 0.3 |
| Grain<sup>8</sup> | 9.7 (0.3) | 15.2 (0.6) | 9.6 (0.2) | 14.3 (0.4) | 8.8 (0.1) | 12.1 (0.2) | <0.0001* | <0.0001* |
| Snack/Sweet<sup>9</sup> | 8.5 (0.4) | 13.8 (0.8) | 9.5 (0.2) | 15.4 (0.3) | 10.2 (0.1) | 15.1 (0.2) | <0.0001* | 0.2 |
| Fruit<sup>10</sup> | 4.6 (0.3) | 2.8 (0.2) | 4.7 (0.2) | 2.7 (0.1) | 4.6 (0.1) | 2.5 (0.1) | 0.9 | 0.05 |
| Vegetable<sup>11</sup> | 11.0 (0.6) | 6.1 (0.5) | 11.0 (0.3) | 6.3 (0.2) | 10.4 (0.1) | 5.4 (0.1) | 0.1 | 0.0009* |
| Beverage<sup>12</sup> | 14.8 (0.3) | 6.1 (0.4) | 13.9 (0.3) | 7.3 (0.3) | 13.8 (0.1) | 9.5 (0.2) | 0.09 | <0.0001* |
| Alcohol<sup>13</sup> | 0.9 (0.2) | 2.4 (0.5) | 1.2 (0.1) | 2.9 (0.3) | 2.1 (0.1) | 5.0 (0.2) | <0.0001* | <0.0001* |
| Water<sup>14</sup> | 10.2 (0.4) | 0.0 (0.0) | 10.1 (0.4) | 0.0 (0.0) | 10.2 (0.2) | 0.1 (0.0) | 0.9 | 0.0011* |
| Fat/Oil<sup>15</sup> | 7.1 (0.4) | 4.0 (0.3) | 6.6 (0.2) | 3.5 (0.1) | 6.1 (0.1) | 3.2 (0.1) | 0.002 | 0.003 |
| Cond<br><sup>16</sup>/Sauce<sup>17</sup> | 4.2 (0.3) | 0.9 (0.1) | 4.2 (0.2) | 0.9 (0.1) | 4.6 (0.1) | 1.0 (0.0) | 0.09 | 0.6 |
| Sugars<sup>18</sup> | 3.8 (0.3) | 0.8 (0.1) | 3.8 (0.2) | 1.1 (0.1) | 3.4 (0.1) | 1.2 (0.0) | 0.05 | 0.08 |
| Other | 0.2 (0.1) | 0.1 (0.0) | 0.2 (0.0) | 0.1 (0.0) | 0.4 (0.0) | 0.3 (0.0) | 0.006 | <0.0001* |

<sup>1</sup> The What We Eat in America broad food categories were applied to categorize all foods and beverages reported in a single day to 14 broad food groups;
<sup>2</sup> Survey weights and adjustments for the complex survey design were applied to represent the non-institutionalized U.S. population. Total numbers and percentages do not always add up to sample size due to missing values and rounding;
<sup>3</sup> Wtd % stands for the estimated weighted percent of all reports of foods or beverages or energy from reported foods or beverages reported in a single day that are included in a food group;
<sup>4</sup> p-values were calculated using the Rao – Scott modified chi-square statistic; “\*” indicates $p < 0.05/42$ or $p < 0.0012$ using a Bonferroni type adjustment for multiple comparisons for food group intake among 14 broad food categories × 3 groups;
<sup>5</sup> Milk, flavored milk, dairy drinks and substitutes, cheese and yogurt;
<sup>6</sup> Meats, poultry, seafood, eggs, cured meats/poultry, and plant-based protein foods;
<sup>7</sup> Mixed dishes containing meat, poultry seafood; grain-based; Asian; Mexican; pizza; sandwiches, and soups;

## 4 Discussion

Previously, few studies have investigated the diet of insulin takers (8–10) nor included consideration of intake contributions by both energy and frequency. To our knowledge, this is the first study to investigate and compare the diets of insulin takers, those who have T2D but do not use insulin, and participants without diabetes. Participants with varying diabetes and insulin using status showed significant differences in both the frequency of which certain foods or beverages are consumed and the energy contributions attributed to certain foods or beverages, implying that insulin using status may be linked to dietary behaviors and the importance of certain foods or beverages in the overall diet. In some broad WWEIA food categories, the statistically significantly differences observed in the frequency were also observed in energy contribution. Both consumption frequency and energy contribution of grains (wheat bread) and alcoholic beverages (beer) were statistically significantly different among the three groups.

The results also included statistically significant findings where either frequency or energy contribution differed among the groups but not both. Beverages, water, and the “other” (nutrition powder) broad food categories did not exhibit significantly different frequency of intake across the three groups, but the share of energy contributed by these broad food categories was lower among insulin takers compared to participants without diabetes. The share of energy contributed by protein (eggs and omelets, cold cuts and cured meats) and vegetables (tomatoes and lettuce) was also greater among insulin takers compared with participants without diabetes. Snacks / Sweets had lower consumption frequency among insulin takers compared with participants without diabetes, but the energy intake was not significantly different across all three groups.

In this study, insulin takers exhibited a pattern of “higher diet quality” regarding the foods they consumed frequently and that contributed most to energy compared to those with T2D not using insulin and participants without diabetes. Since older adults accounted for the majority of the insulin takers, this finding is consistent with previous studies showing that older adults have higher-quality diets (10,23,25,26). In addition, that insulin takers have a higher diet quality is also supported by a previous study (10), suggesting that those with diabetes who are using insulin engage in dietary behaviors promoting a more healthful mix of dietary intake compared with those who do not have diabetes and those with T2D. The group using insulin reported a higher consumption frequency of grains, energy contribution from grains, protein foods and vegetables; and lower consumption frequency of alcohol and sweets/snacks and energy contributed by beverages and alcohol compared to participants without diabetes. One explanation is that grains, protein foods, and vegetables have been promoted as foods to consume to help manage blood glucose control for those taking insulin. Carbohydrate intake is highly related to postprandial glycemia, whole grains are a source of carbohydrates that have a favorable influence on postprandial blood glucose levels (27). Protein foods may also aid dietary compliance and weight loss maintenance, which is beneficial for diabetes (28,29). Vegetables consumption has been associated with the improvement of diabetes due to benefits offered by the bioactive compounds comprising them including dietary fiber, resistant starch, antioxidant vitamins, and minerals (30–32), which may explain why insulin takers select a higher percentage of their calories from vegetables compared with those who do not have diabetes. Furthermore, alcohol, sweets/snacks, and beverages (heavily influenced by sweetened beverages) are foods that those taking insulin may try to avoid due to their negative impact on glycemic control. These foods that may add solid fats or added sugars and intake among all Americans should be reduced according to the Dietary Guidelines for American, 2020-2025 (33,34). Previous studies found that those with diabetes drink less alcohol compared to those without diabetes (35,36), which supports the findings here that those who used insulin and those with T2D not taking insulin, consumed less alcohol than participants without diabetes. Even though the long-term effect of alcohol consumption on glycemic management remains to be investigated (37), heavy alcohol consumption is associated with an increased incidence of diabetes (37) and alcohol consumption may be a negative indicator of diabetes self-care behavior (38). The findings related to lower frequency of sweets/snacks and beverages among those using insulin compared to those without diabetes is supported by a previous study where participants with diabetes consumed lower amounts of sweets and beverage (juice) compared to those without diabetes (36). Sweets/snacks and beverages especially sweetened beverages that heavily add to energy intake, are typically high in added sugar. Added sugar intake is not only a significant contributor to weight gain that can lead to increased risk of diabetes, but also increased dietary glycemic load and fructose metabolism that leads to inflammation and insulin resistance (39). However, it should be noted that the literature reviewed here includes studies focused on those with diabetes, not specifically those using insulin, due to the extremely limited evidence of dietary intake among those using insulin (31,32,36). Therefore, the findings here should be further explored among those taking insulin.

Limitations of this analysis may be due to the aggregation of foods to subcategories and categories that may obscure specific food items or types of food that could be responsible for much of the differences among groups. The addition or removal of specific foods or beverages from a food category has the potential to change statistically significant comparisons so the results and interpretation should be viewed as one perspective on the dietary intake among these three groups keeping in mind that analysis of alternative food categorization, splitting to subcategories or itemization may yield differential results. However, the application of the WWEIA food categories lends standardization and consistency with other reporting by USDA and other researchers (40–42). In addition, the ranking lists (table 2-7) were generated from the foods and beverages reported over all times of the day, so separating out the most frequently consumed foods and foods that highly contributed to energy intake for different times or meals of the day may be helpful to inform interventions or dosing algorithms as the timing of dietary intake and combinations of foods consumed at the same time influences glucose metabolism (43–47). Furthermore, the NHANES dietary data this study used is only a one-day dietary recall of the participants, which may not be accurate due to participants’ memory and may not represent the usual dietary intake (48). Moreover, misreported dietary intake may have been presented in this study sample resulting in errors in energy, frequency, food item, subcategory, and category representation and comparisons (49). Those with diabetes and insulin takers may exhibit different patterns of misreporting compared with other population groups (36). Previous studies have shown that dietary intake misreporting and especially underreporting is more prevalent in overweight and obese individuals compared to normal weight individuals (50–52), and prevalence of underreporting in diabetic patients may be even higher (53), considering that obesity is a risk factor of diabetes that 81% of diabetic patients are obese (54). Another previous study pointed out that energy misreports are prone to misreport foods that highly contributed to glycemic index and glycemic load such as underreporting foods including sugars, cookies, and milk products and overreporting foods including fruit and vegetables (52). For future study, the differences in misreporting by diabetic status should be evaluated.

There are several implications and applications for the results of this study. Diabetes is a chronic disease that is highly related to diet (55). The lists of foods generated in this study may be used to fill the gap in knowledge of the diets of U.S. adult insulin takers or diabetic patients (8,9), specifically, menus and dietary interventions developed to improve diabetes management may be informed by the lists. Although frequently consumed foods and those contributing most to energy may not satisfy the preferences of every person, they are generally accepted as indicated by their selection in the population. Those using insulin may be more likely to adhere to a diet they accept, thus forming healthier eating habits. Second, the findings of this study may inform future studies to gain a greater knowledge of the diets of those using insulin. For example, the list of frequently consumed foods and those most contributing to energy may inform the prioritization of foods and beverages to include in food frequency questionnaires. Third, the findings of this study may aid the development of nutrition assessment tools as researchers should concentrate efforts on developing the most accurate identification methods for the foods and beverages that are consumed the most frequently and contribute the most to total energy intake in the group they are tailored for (23).

## 5 Conclusions

Differences in dietary intake exist among U.S. adults by diabetic status using NHANES 2009-2016. Insulin takers consumed grains more frequently, while consuming snack/sweet and alcohol less frequently compared to participants without diabetes. Protein, grains, and vegetables contributed more to insulin takers’ daily total energy intake compared to participants without diabetes, whereas beverage and alcohol contributed less. Frequently consumed foods and those contributing to energy may inform diabetes control and management, tailored food recipe, consumer education, questionnaire design, and dietary assessment.

## Data Availability

All data produced in the present study are available upon reasonable request to the authors

https://www.cdc.gov/nchs/nhanes/index.htm

## 7 Author Contributions

HAE-M and LL designed research. LL analyzed data and drafted the manuscript. HAE-M, FZ, and ED critically reviewed and edited the manuscript. HAE-M had primary responsibility for final content. All authors read and approved the final manuscript.

